# Genome-wide association studies found *CCDC7* and *ITGB1* associated with diabetic retinopathy

**DOI:** 10.1101/2024.08.10.24311791

**Authors:** Tengda Cai, Qi Pan, Yiwen Tao, Luning Yang, Charvi Nangia, Aravind L Rajendrakumar, Yu Huang, Yongqing Shao, Yunyan Ye, Tania Dottorini, Mainul Haque, Colin NA Palmer, Weihua Meng

## Abstract

**Purpose:** Diabetic retinopathy (DR), a complication affecting the eyes, is associated with diabetes. This study aims to identify genetic variants associated with DR in patients with type 1 diabetes in the UK Biobank cohort (n = 1,004).

**Methods:** A genome-wide association study (GWAS) was conducted to identify significant genetic variants of DR in type 1 diabetes. The findings are set to undergo validation during the replication and meta-analysis stages by using six cohorts: African American, European, FinnGen, GoSHARE, GoDARTS and Caucasian Australians.

**Results:** In a locus, top single nucleotide polymorphism (SNP) rs184619214 in *CCDC7* reached a GWAS significance level (*p* = 6.38 x 10^−9^) and rs79853754 in *ITGB1* (*p* = 3.24 x 10^−8^), with both genes being adjacent to each other. The SNP-based heritability was estimated to be 31.09%. Rs184619214 was replicated and reached statistical significance (*p* < 5.0 x 10^−8^) in the meta-analysis stage. Pathway analysis revealed that ITGB1 is involved in the generation of biomolecules that impact the progression of DR. PheWAS analysis revealed that osteoarthritis (OA) of the hip was significantly associated with most of the SNPs of the locus. Mendelian Randomization further confirmed an association between OA and DR.

**Conclusions:** Our study has identified a novel genomic risk locus associated with DR in type 1 diabetes, located in the intergenic region between the *CCDC7* and *ITGB1* genes, providing insights for DR researchers.

## Introduction

Diabetic retinopathy (DR), a major microvascular complication of diabetes mellitus (DM), is a leading cause of vision loss due to diabetes-induced retinal damage. Research suggests that among individuals with diabetes, the global occurrence of DR is estimated to be 34.6%, 6.96% for proliferative diabetic retinopathy (PDR) ^1^. The incidence of DR increases over time, affecting up to 50% of patients within 10 years of a diabetes diagnosis, and 90% by 25 years ^2^. There are well-described risk factors for the development of DR. The most clearly recognized are the duration of diabetes (DoD) and the degree of glycaemic control measured as glycosylated hemoglobin (HbA1c) ^3^. The burden of DR is anticipated to grow due to factors such as an aging population, increasing obesity rates, and the expanding DM epidemic.

In patients with glycated hemoglobin (HbA1c) ≤ 7.0%, the prevalence of any type of DR was 18.0%, compared with 51.2% in patients with HbA1c > 9.0% ^4^. The Wisconsin Epidemiological Study of Diabetic Retinopathy (WESDR) shows that DM duration is a key predictor of DR progression, with DR prevalence increasing from 17% in type 1 DM and 29% in type 2 DM for those with less than 5 years of DM, to nearly 100% in type 1 DM and 78% in type 2 DM for those with the disease for over 15 years ^5^. This trend is further supported by a study showing that the prevalence of DR escalates with the DoD, with 21.1% of patients affected at less than 10 years, soaring to 76.3% at 20 years or more ^6^. Studies suggest that there is a clear familial nature to DR, with twin and family studies suggesting a genetic basis, yet the specific genes contributing to DR risk remain elusive, with only a handful of genetic associations identified, none of which demonstrate strong links ^5,7^.

In a study of 4,172 patients diagnosed with T1D from 12 years of age, 26.7% had background diabetic retinopathy, 10.7% had initial diabetic retinopathy, and 4.1% had PDR ^8^. Notably, adults with T1D indicated that the prevalence of DR increases with the disease duration, affecting more than 80% of patients who have lived with the disease for over 40 years ^9^. In the United States, T1D affects 244,000 youths and 1.6 million adults over 20 years old, with around 64,000 new cases diagnosed annually ^10^. This means that cases of DR caused solely by T1D are not uncommon. T1D itself results from the immune-mediated destruction of insulin-secreting beta cells in the pancreas, leading to chronic hyperglycemia and subsequent organ impairment, including the eyes ^8,11^. Alarmingly, the incidence of T1D and related vision loss is on the rise, growing annually at a rate of 3% to 5% ^12^. Effective interventions such as strict glucose and blood pressure management, lipid-lowering treatments, and laser therapies can slow disease progression and help preserve vision ^13^.

Various genome-wide association studies (GWAS) have identified numerous genetic loci potentially linked to DR, highlighting a significant genetic component in its pathology ^13,14,15,16^. Specifically, for T1D-related DR, GWAS have pinpointed candidate genes such as *MIR449b* ^11^, *CACNB2* ^17^, and others including *LOC339529, RBFOX1* ^18^, *KLRD1*, *TNF* ^19^, as well as *KIF16B, FUT4* ^20^. However, these identified GWAS loci for DR often vary and lack consistent replicability across studies, which complicates deeper genetic analyses and hinders the identification of novel biological pathways for targeted interventions. This gap in understanding exacerbates the healthcare challenges and costs associated with managing DR.

To elucidate the genetic mechanisms linked to DR in T1D patients, we conducted a GWAS based on the UK Biobank (UKB) cohort, with efforts using additional cohorts for replication. This research aims to uncover new genetic loci related to DR, providing valuable information for researchers in this area.

## Research Design and Methods

### Information on Cohorts and Patients

The UKB functions as a large-scale biomedical database and research resource, housing detailed genetic and phenotypic genetic and phenotypic information for approximately 500,000 participants throughout England, Scotland, and Wales. For further information on the UKB cohort, refer to www.ukbiobank.ac.uk. Conducted in accordance with the Declaration of Helsinki’s tenets, this study secured informed consent from all participants after they were fully informed about the UKB study’s purpose. The research received the necessary ethical approval from the National Health Service National Research Ethics Service (reference 11/NW/0382).

In this research, the genetic data came from the UK Biobank cohort. They used a standardized process for DNA extraction and quality control (QC), in which detailed methods can be found at https://biobank.ctsu.ox.ac.uk/crystal/ukb/docs/genotyping_sample_workflow.pdf. We defined samples clearly for cases and controls by phenotype data filtering using the following UKB field ID codes (Fig.1). Our study used the participants of white British (UKB field ID: 21000). T1D with prolonged DoD was identified using four criteria: a diagnosis of diabetes by doctors (UKB field ID: 2443), a response of ‘Yes’ to the question ‘Started insulin within one year of diabetes diagnosis’ (UKB field ID: 2986), “age diabetes diagnosed” <= 40 years old (UKB field ID: 2976), and diabetes samples that have a DoD at least 20 years. Among T1D patients, DR cases were identified using the ICD-10 code ‘H360’ from the World Health Organization International Classification of Diseases (UKB field ID: 41202). Conversely, control subjects were those without the ICD-10 code ‘H360’, indicating no diagnosis of DR. Furthermore, we selected the HbA1c level that ranks in the top 95% of the above-selected samples to focus on individuals with relatively poor glycemic control, which is a critical risk factor for the development of DR. The clinical characteristics of the case and control groups are shown in Table 1. Six cohorts were used for replication and meta-analysis, including African American & European cohorts^16^, FinnGen cohorts ^21^, GoSHARE & GoDARTS (Rajendrakumar, 2022), Caucasian Australians^22^. More information is shown in Supplementary Table S1.

**Fig. 1.**
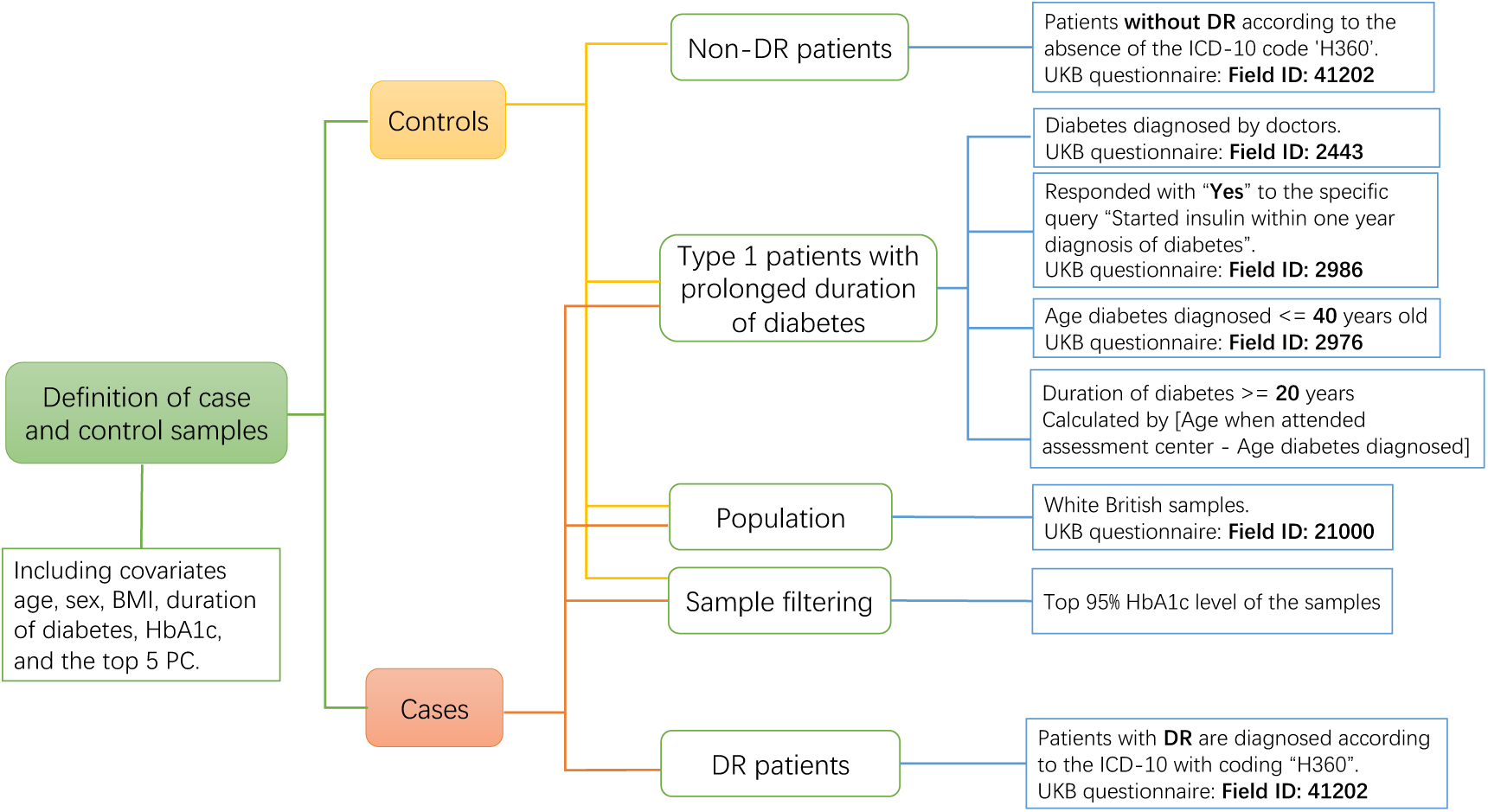
Definition of diabetic retinopathy in type 1 diabetes based on UKB cohort. The control samples consist of non-DR patients, T1D with prolonged duration of diabetes, white British ethnicity, and those within the top 95% of HbA1c levels. The case samples include DR patients, T1D with prolonged duration of diabetes, white British ethnicity, and those within the top 95% of HbA1c levels. The GWAS model incorporates covariates age, sex, BMI, duration of diabetes, HbA1c levels, and the top 5 principal components.

**Table 1.** Clinical characteristics using a dataset based on UK Biobank.

| Clinical Characteristics |  |  |  |
| --- | --- | --- | --- |
| Covariates | Cases | Controls | *P* |
| Sex(male:female) | 256:192(57.14%:42.86%) | 315:241(56.65%:43.35%) | 0.87 > 0.05 |
| Age(years) | 57.1(8.02) | 55.1(7.66) | $5.81 \times 10^{-5} < 0.05$ |
| BMI(kg/m<sup>2</sup>) | 27.7(4.58) | 27.0(4.59) | 0.03 < 0.05 |
| DoD(duration) | 36.4(9.74) | 33.7(9.29) | $1.05 \times 10^{-5} < 0.05$ |
| HbA1c(mmol/mol) | 64.9(13.25) | 60.7(10.98) | $1.15 \times 10^{-7} < 0.05$ |

DoD: duration of diabetes. A chi-square test assessed the differences in sex distribution between cases and controls, while an independent t-test evaluated other covariates. Continuous covariates were shown as 'mean(standard deviation)'.

### Statistical analysis

Genome-Wide Complex Trait Analysis (GCTA v1.93.3) was used for GWAS association analysis (refer to https://yanglab.westlake.edu.cn/software/gcta/#Overview). Our study performed the genome-wide association methods utilizing the fastGWA-GLMM tool ^23^. Covariates incorporated into the model were age, sex, BMI, DoD, HbA1c, and the top 5 genetic principal components. In the QC step, SNPs with imputation INFO scores < 0.3 and minor allele frequency < 0.5% were removed by using GCTA, as well as SNPs that failed Hardy-Weinberg < 0.00001 and samples with missing call rates < 0.1 were filtered out by using Plink v1.90. SNPs situated on the X and Y chromosomes and mitochondrial SNPs were eliminated from consideration. SNPs were considered to reach genome-wide association significance if they had a *p* value less than 5×10^−8^. SNP-based heritability was calculated using the “BLD-LDAK Model” in SumHer ^24,25^.

### Replication studies and Meta-analysis

Our replication approach is to replicate SNPs or their LD proxies in other studies that matched the significant SNPs (p < 5×10^−8^) found in our GWAS and record their p values. Fixed-effects meta-analyses with inverse weighting were conducted using the R package ‘metafor’. Notably, to standardize and align the strand orientation for alleles across different cohorts, we have inverted the strand designation (switching the effect allele, EA, with the non-effect allele, NEA) in summary statistics results produced from our UKB study. Correspondingly, the beta values and their 95% confidence intervals have also been adjusted to ensure consistency in the results.

### GWAS analysis by FUMA and LDSC

The GRCh37 genome assembly was followed for SNP annotation. SNP functional annotations and Manhattan plots were applied and produced using FUMA v1.5.2, a web platform designed to annotate, prioritize, visualize, and interpret GWAS results ^26^. Gene mapping was conducted for cis-expression quantitative trait loci (cis-eQTL), chromatin interaction, and positional mapping studies. Gene expression heatmaps were generated, and tissue specificity analyses were performed for genes identified through positional mapping, utilizing data from 54 specific tissue types and 30 general tissue types in GTEx v8. The gene analysis, gene-set analysis, and tissue expression analysis were performed by MAGMA v1.08, which was integrated into FUMA. In gene analysis, SNP summary statistics were consolidated at the gene level, assessing the combined association of all SNPs within a gene with the phenotype. Gene-set analysis involved grouping genes based on common biological, functional, or other traits, which helped to unravel the role of specific biological pathways or cellular functions in the genetic basis of a phenotype. To test the relationship between highly expressed genes in a specific tissue and genetic associations with DR, tissue expression analysis, based on GTEx (https://www.gtexportal.org/home/), was performed separately for 30 general tissue types and 53 specific tissue types. The parameters in the FUMA platform are set in Supplementary Table S2. Q-Q plots and regional visualization were provided by LocusZoom (http://locuszoom.org/).

### Enrichment of genes and Pathway analysis

Enrichment analysis of positional gene sets was performed using the ‘GENE2FUNC’ feature in FUMA. Utilizing the Molecular Signatures Database (MSigDB) ^27,28^ and the Reactome ^29^, we identified relevant pathways and gene interactions for genes of interest, particularly *ITGB1* and *CCDC7*. This analysis involved mapping our significantly associated SNPs to corresponding gene sets in MSigDB and exploring gene interactions through Reactome pathways to assess the potential biological mechanisms influencing diabetic retinopathy.

### Transcriptome-wide association studies (TWAS)

We employed Transcriptome-Wide Association Studies (TWAS) (http://gusevlab.org/projects/fusion/) ^30^ to assess the impact of gene expression modulated by genetic variants on DR risk. Focusing on specific tissues from panel GTEx v7, TWAS integrated SNP-gene expression associations from eQTL studies with SNP-disease associations from GWAS summary statistics.

### Phenome-Wide Association Study

To investigate potential associations between the top SNPs identified in our current analysis and other related traits, we conducted a Phenome-Wide Association Study (PheWAS). In this analysis, a PheWAS was performed using a comprehensive dataset of 4,756 GWAS summary statistics available on the GWASATLAS platform (https://atlas.ctglab.nl/PheWAS). Our inclusion criteria included all GWAS. To construct the PheWAS SNP heatmap, we only included phenotypes that had p values less than 0.05 in their association with the SNPs. Construct a data frame with 12 SNPs associated with different traits. Adjustment for multiple comparisons was performed using the Bonferroni correction method.

### Mendelian Randomization

To investigate whether significant phenotypic results from PheWAS are causally related to DR, we used a bidirectional Mendelian randomization (MR) approach using the “TwoSampleMR” package. Except for DR summary statistics, data sources were obtained from the IEU OpenGWAS database (https://gwas.mrcieu.ac.uk/). Prior to MR analysis, the data were clumped to address the issue of linkage disequilibrium. Our MR methods included inverse variance weighted (IVW), MR Egger, weighted median, simple median, and weighted mode.

## Results

### Significant GWAS results

We identified 448 cases and 556 controls after genetic and phenotype quality control. Altogether, 11,185,683 imputed SNPs passed from routine QC checking and imputation INFO quality scores that were larger than 0.3. The most significant SNP in this region was rs184619214 in the *CCDC7* gene with a *p* value of 6.38 x 10^−9^ in Chromosome 10 (see Table 2). The significant SNP rs79853754 in *ITGB1*, with a *p* value of 3.24 x 10^−8^, is in high linkage disequilibrium with rs184619214 (r^2^ = 0.95). A cluster of top SNPs reached a significant level in the Manhattan plot, with regional plots at the top (Fig.2). The corresponding Q-Q plot is shown in Fig.3. The SNP-based heritability of DR was estimated to be 31.09% in this T1D population.

**Fig. 2.**
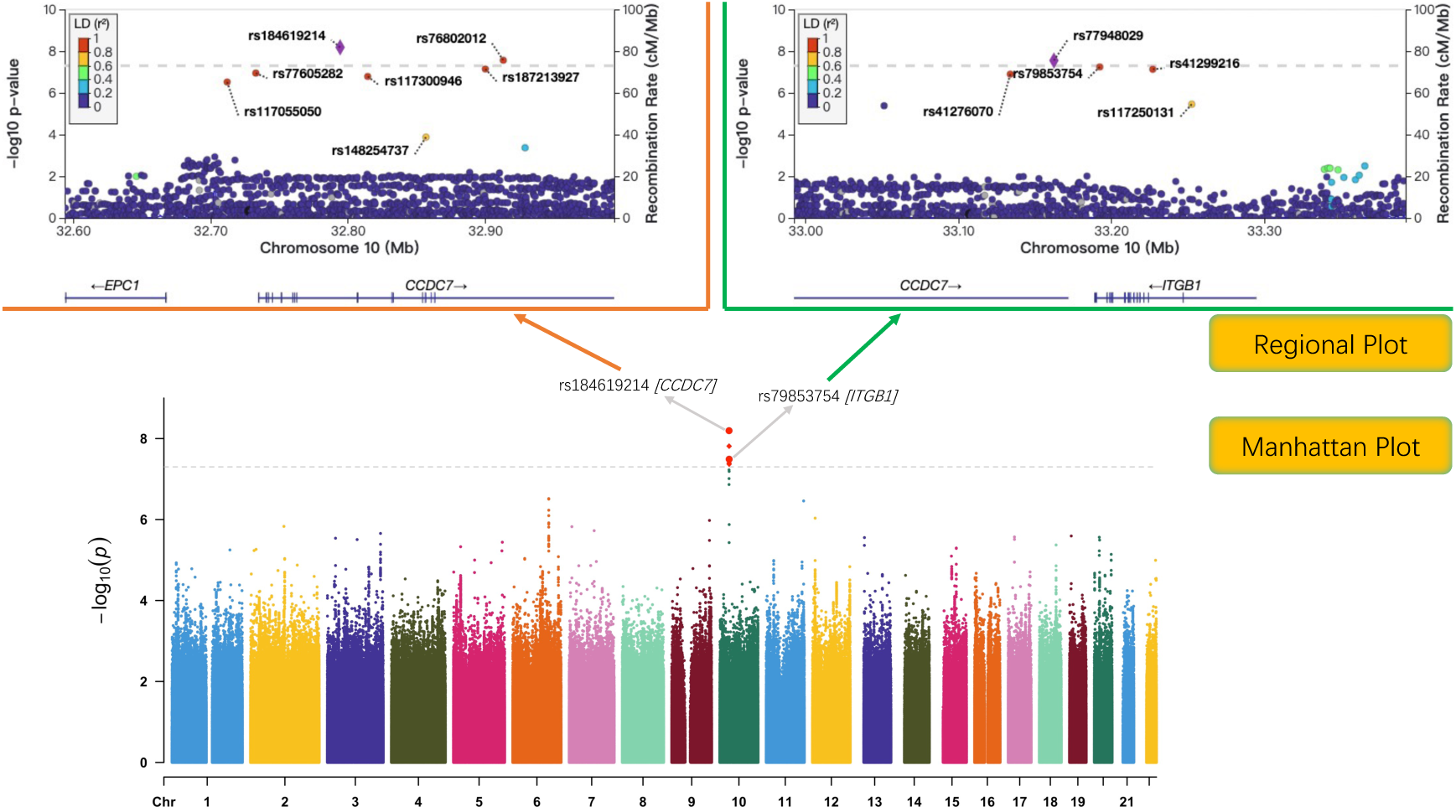
The Manhattan and regional plots of the GWAS analysis for diabetic retinopathy in type 1 diabetes. Figure below displays a cluster of top SNPs that reached a significant level in the Manhattan plot, with regional plots positioned above it. The regional plot on the left primarily includes SNPs within the *CCDC7* gene, while the regional plot on the right predominantly features SNPs within the *ITGB1* gene.

**Fig. 3.**
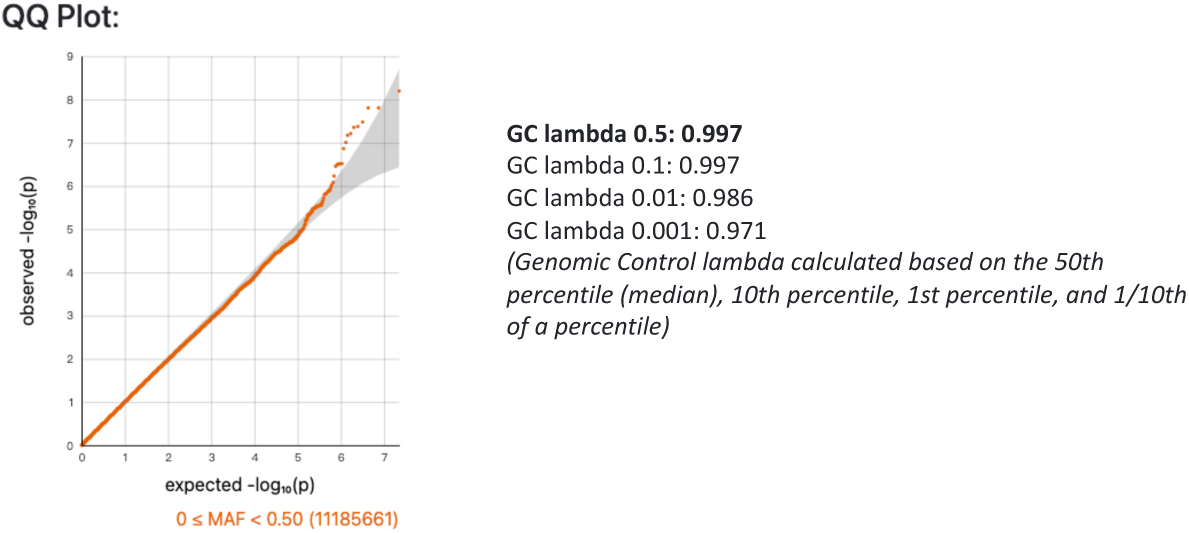
The Q-Q plot of GWAS for DR in type 1 diabetes. This QQ plot visually assesses the distribution of observed p-values (plotted on the y-axis) against the expected distribution under the null hypothesis (plotted on the x-axis) for the GWAS on DR. The linearity of the points along the diagonal suggests that the observed p-values largely conform to the expected distribution, indicating well-controlled population stratification with genomic control lambda value being very close to 1.

**Table 2.** Significant locus of GWAS based on the UKB.

| Marker | Chr | Position | Nearest gene | EA | NEA | AF1 | Beta | SE | $p$ value | $r^2$ |
| --- | --- | --- | --- | --- | --- | --- | --- | --- | --- | --- |
| rs184619214 | 10 | 32794461 | *CCDC7* | A | G | 0.98 | -0.47 | 0.08 | 6.38E-09 | 1.00 |
| rs76802012 | 10 | 32913261 | *CCDC7* | A | T | 0.98 | -0.45 | 0.08 | 1.53E-08 | 0.96 |
| rs77948029 | 10 | 33162616 | *CCDC7* | A | T | 0.98 | -0.45 | 0.08 | 1.55E-08 | 0.96 |
| rs79853754 | 10 | 33192606 | *ITGB1* | G | C | 0.98 | -0.44 | 0.08 | 3.24E-08 | 0.95 |
| rs187213927 | 10 | 32900213 | *CCDC7* | A | G | 0.98 | -0.45 | 0.08 | 4.04E-08 | 0.95 |
| rs41299216 | 10 | 33227260 | *ITGB1* | G | A | 0.98 | -0.45 | 0.08 | 4.29E-08 | 0.93 |
| rs77605282 | 10 | 32733099 | *CCDC7* | C | T | 0.98 | -0.45 | 0.08 | 5.93E-08 | 0.97 |
| rs41276070 | 10 | 33134064 | *CCDC7* | A | G | 0.98 | -0.45 | 0.08 | 6.48E-08 | 0.95 |
| rs117300946 | 10 | 32814555 | *CCDC7* | A | G | 0.98 | -0.43 | 0.08 | 9.73E-08 | 0.92 |
| rs117055050 | 10 | 32712210 | *CCDC7* | G | A | 0.98 | -0.48 | 0.09 | 1.37E-07 | 0.91 |
| rs117250131 | 10 | 33252757 | *ITGB1-DT* | A | C | 0.98 | -0.36 | 0.07 | 1.33E-06 | 0.89 |

\* indicates SNPs reach genome-wide significance with $p$ value $< 5 \times 10^{-8}$ , while others are suggestive.
Chr: chromosome, EA: effect allele, NEA: non effect allele, AF1: frequency of effect allele

### Replication & Meta-analyses

Rs184619214 was replicated in the African American and European cohorts with a *p* value of 0.05 and 0.04. Meta-analyses conducted with six cohorts revealed that rs184619214 reached genome-wide significance with *p* value of 4.47 x 10^−11^ (Table 3). Figure S1 shows a forest plot, funnel plot and radial plot of meta-analysis results using UKB, European, FinnGen, GoSHARE, GoDARTS, and Caucasian Australian cohorts.

**Table 3.** *p* values of top significant SNP rs184619214 from UKB GWAS in replication cohorts and meta-analysis results ^16,21,23, (Rajendrakumar, 2022)^

| GWAS | Replication results for significant SNP rs184619214 from UKB |  |  |  |  |  |  |  |  |  | Meta-analysis results |  |  |  |  |
| --- | --- | --- | --- | --- | --- | --- | --- | --- | --- | --- | --- | --- | --- | --- | --- |
|  | SNPs(LDproxies) | CHR | POS | EA | NEA | AF1 | BETA | SE | *P* | DISTANCE | ESTIMATE | SE | *P* | Q_heterogeneity | Q_Pval |
| African American | rs139204448 | 10 | 31864160 | C | T | 0.017 | / | / | 0.050 | -930,301 | 0.329 | 0.05 | 4.47E-11 | 8.91 | 0.11 |
| European | rs78963187 | 10 | 32073079 | G | T | 0.035 | 0.453 | 0.13 | 0.040 | -721,382 |  |  |  |  |  |
| FinnGen | rs184619214 | 10 | 32505533 | G | A | 0.054 | 0.120 | 0.31 | 0.699 | 0 |  |  |  |  |  |
| GoSHARE | rs11008927 | 10 | 32703688 | A | C | 0.250 | 0.126 | 0.16 | 0.436 | -90,773 |  |  |  |  |  |
| GoDARTS | rs17230291 | 10 | 32646193 | G | A | 0.036 | 0.168 | 0.10 | 0.095 | -148,268 |  |  |  |  |  |
| Caucasian Australian | rs2488320 | 10 | 33238917 | T | C | 0.354 | 0.245 | 0.17 | 0.150 | 404,450 |  |  |  |  |  |

"/" indicates missing value. Chr: chromosome, EA: effect allele, NEA: non-effect allele, AF1: frequency of effect allele.
The effect alleles and beta values of the SNPs in the UKB were adjusted to ensure that the strand direction was consistent across all cohorts during meta-analysis.

### Gene, gene-set and tissue expression analysis by MAGMA

The entire dataset of GWAS summary statistics was utilized to conduct a comprehensive assessment of gene-disease associations.

In the gene analysis, all the SNPs located within genes were mapped to 19,295 protein-coding genes. Genome-wide significance was defined (red dashed line in the plot) at *p* = 0.05/19295 = 2.59 x10^−6^. No gene demonstrated the strongest association with DR, see Figure S2.

In the gene-set analysis, 10,678 gene sets from MsigDB v6.2 were tested. The top first gene set GOMF_LEUKOTRIENE_C4_SYNTHASE_ACTIVITY has not achieved a statistically significant association after Bonferroni correction, demonstrating a *p*-value of 1.61×10^−5^ (> 0.05/10,678 = 4.68×10^−6^) and *p_bon_* = 0.27 < 0.05. This analysis’s top ten gene sets are shown in Supplementary Table S3.

Tissue expression analysis demonstrated no tissue achieved statistical significance in the expression analysis of 30 general tissue types from multiple organs and the 53 specific tissue types within some organs. Results are shown in Figure S3.

### Cis-expression quantitative trait loci (cis-eQTL), chromatin interaction analysis and positional mapping

Cis-eQTL analysis did not identify any significant SNPs associated with GTEx tissue types. Chromatin interaction analysis revealed that significant occurring tissues or cells in *CCDC7* are Left_Ventricle, IMR90, Mesenchymal_Stem_Cell, Mesendoderm, Trophoblast-like_Cell, hESC, and in *ITGB1* are Left_Ventricle, Liver, GM12878, IMR90, Mesenchymal_Stem_Cell, Mesendoderm, Trophoblast-like_Cell, hESC respectively. The results of chromatin interactions are presented in the Fig.4, genes interacting with *CCDC7* and *ITGB1* were *NRP1*, *C10ORF68*, *EPC1*, *ARHGAP12*, *ZEB1*, *ZNF438* and *MAP3K8*. Positional mapping identified 10 genes as potentially related to the DR, including *CCDC7*, *ITGB1*, *NRP1*, *EPC1*, *ARHGAP12*, etc (details are listed in Supplementary Table S4).

**Fig. 4.**
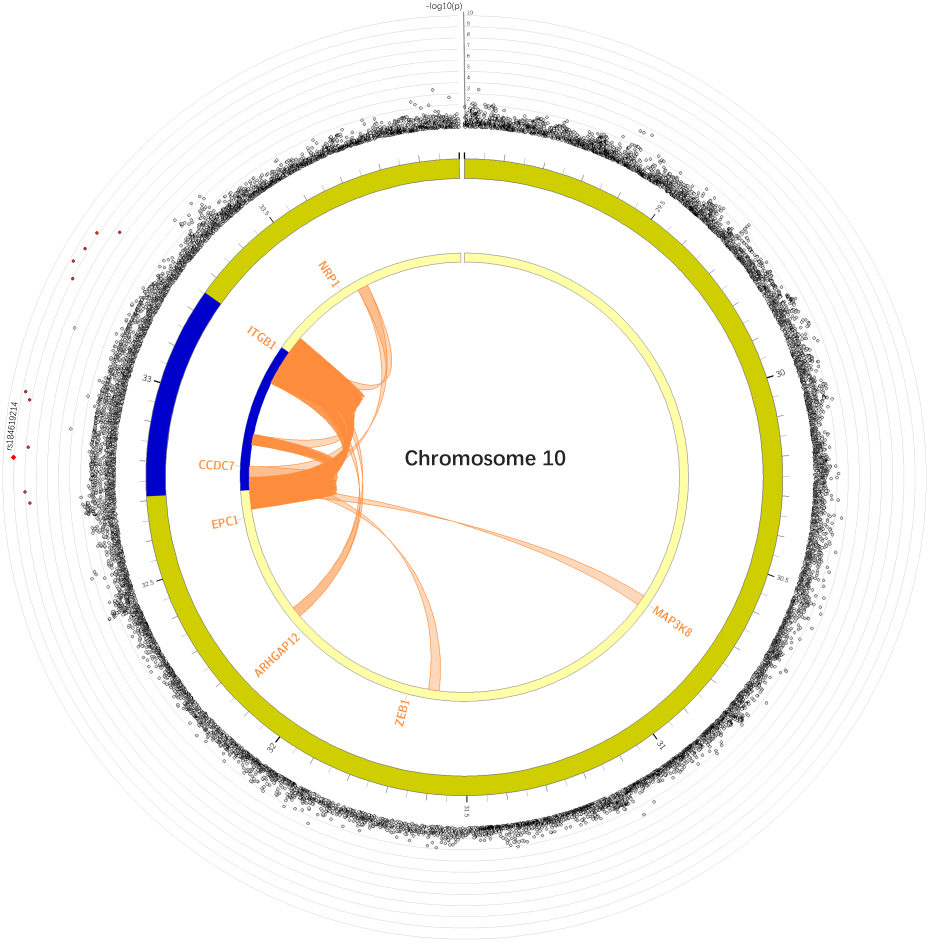
The circus plot of gene mapping of chromatin interactions and cis-eQTL. The top SNPs in each risk locus are displayed in the most outer layer (Manhattan plot). The second and third layers are chromosome rings, where genomic risk loci are highlighted in blue to indicate their locations. Genes mapped exclusively by chromatin interactions are colored orange.

### Gene expression heatmap and tissue specificity

Gene expression heatmaps using average expression value per label are shown in the Fig.5 Cells filled in red represent higher expression compared to cells filled in blue across genes and labels. Cells filled in red represents higher expression of the genes in a corresponding label compared to other labels, but it does not represent higher expression compared to other genes. The expression of gene *CCDC7* in the testis is obviously higher than in other tissue types. Nerve tibial shows a relatively higher expression among all these genes.

**Fig. 5.**
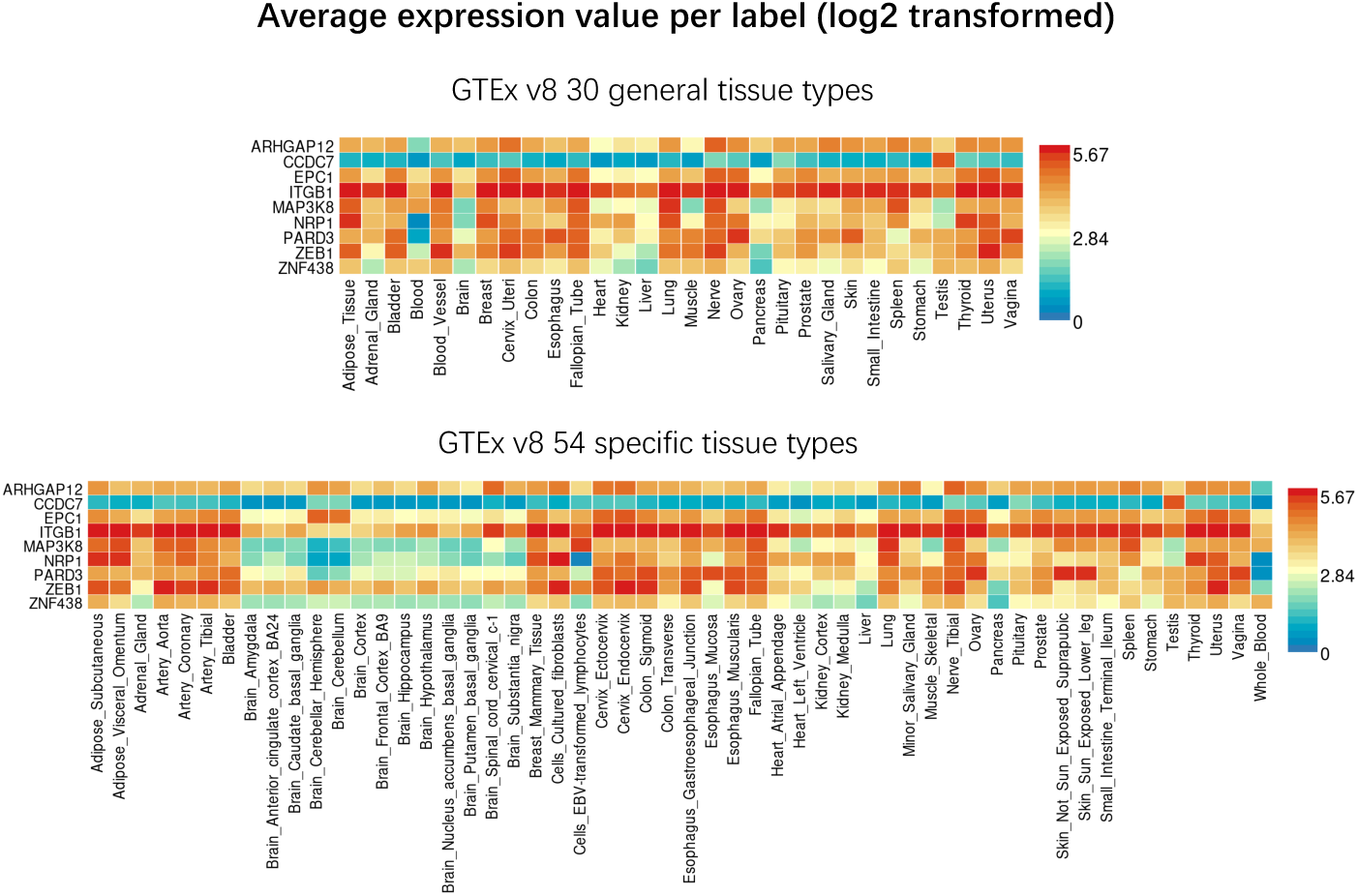
Gene expression heatmaps of positional mapping genes. This figure presents two heatmaps illustrating the average expression values (log2 transformed) of selected positional mapping genes across diverse tissue types, as sourced from the GTEx v8 dataset. The upper heatmap displays gene expression in 30 general tissue types, while the lower heatmap shows expression across 54 specific tissue types. Expression values are color-coded from low (blue) to high (red), offering a detailed view of the tissue-specific expression patterns of these genes.

Tissue specificity is tested using the differentially expressed genes (DEG) defined for each label of each expression data set. An enrichment test of DEG for both GTEx v8 54 tissue types and GTEx v8 30 general tissue types was performed, only testing the enrichment of positional mapping genes instead of using the full distribution of SNP *p* values in MAGMA tissue expression analysis. Tissue specificity testing found that brain cortex showed significant enrichment in the down-regulated DEG in GTEx v8 54 tissue types. See Figure S4.

### Enrichment of genes and Pathway analysis

The results of the gene enrichment are shown in Figure S5. *CCDC7*, *C10ORF68*, *ITGB1* and *NRP1* are overlapping genes. The pathway analysis revealed enrichment and interactions involving *ITGB1* and *CCDC7*. Utilizing the Human Tissue Compendium from MSigDB, we observed associations among these genes and also identified the varying strengths of connections between these genes and various tissues (Figure S6). In the Reactome pathway diagram (Figure S7), *L1CAM* is involved in the assembly of integrin complexes and suggests that *CHL1* mediates interactions between *ITGB1* and *NRP1*.

### TWAS results

TWAS results across multiple tissue types using GTEx v7 data did not reveal statistically significant associations for the SNPs associated with our GWAS results (Supplementary Table S5). Despite extensive testing, both the *CCDC7* and *ITGB1* genes did not show significant expression-trait associations in the tissues analysed. Specifically, *CCDC7* was occasionally identified but not consistently across different tissues, while *ITGB1* was rarely detected, which may indicate a lack of statistical power or the broader biological roles of the genes not limited to tissue-specific expression.

### Phenome-Wide Association Study (PheWAS)

Totally 8 potentially significant phenotypes were selected according to their high association with the 12 SNPs (upper part of Fig.6), including eotaxin (CCL11), fresh fruit intake, M23 internal derangement of knee, osteoarthritis of hip, posterior corona radiata fractional anisotropy, posterior thalamic radiation (include optic radiation) fractional anisotropy, posterior thalamic radiation (include optic radiation) radial diusivities, sagittal stratum fractional anisotropy, as indicated by red coloring predominantly in the heatmap (p < 0.01). After Bonferroni correction, where each SNP has its own threshold (lower part of Fig.6), we found that three phenotypes Posterior thalamic radiation (include optic radiation) fractional anisotropy, Posterior thalamic radiation (include optic radiation) radial diusivities, Osteoarthritis of hip (hospital diagnosed) were significant with most of the SNPs.

**Fig. 6.**
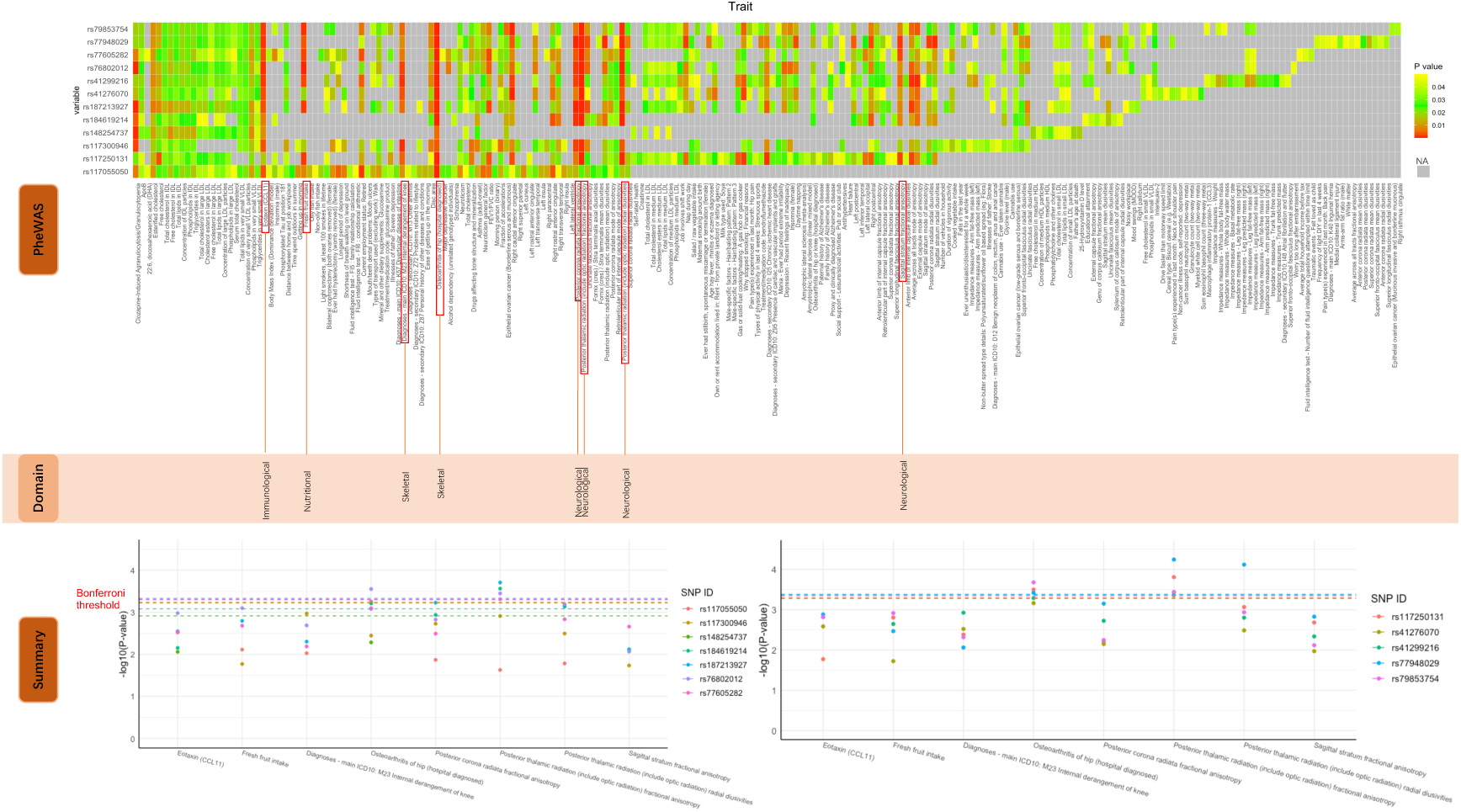
Results of PheWAS for the top 12 SNPs in significant GWAS. This figure presents a multi-part analysis of 12 top SNPs in high LD and their phenome-wide association study results. The top figure (heatmap) displays the pheWAS results, where each cell represents the association of SNPs with various traits, color-coded by p-value significance. The middle panel, labeled ‘Domain,’ highlights the phenotypes concentrated across multiple SNPs (marked in red) and categorizes them into their respective domains. The bottom figure is divided into two sections based on the regional SNP locations from the regional plot. Each section shows the significance of individual SNPs, with dashed lines representing the Bonferroni correction thresholds for each SNP.

### Mendelian Randomization

The results revealed that osteoarthritis (OA) (data from IEU ID: “ebi-a-GCST90038686”), when considered as an exposure, significantly led to DR, with an IVW *p* value of 0.02, which is below the significant threshold of 0.05. In the heterogeneity analysis, both the IVW and MR Egger p-values are greater than 0.05, indicating no significant heterogeneity between the genetic tools used, suggesting consistency in the magnitude and direction of their effects. Conversely, DR as an exposure did not significantly lead to OA, as indicated by an IVW *p* value of 0.10 > 0.05. More detailed results can be found in Supplementary Table S6. The accompanying figure illustrates the MR analysis, including methods: IVW, MR Egger, simple median, weighted median and weighted mode. It plots the SNP effects on osteoarthritis against their effects on diabetic retinopathy in Fig. 7 and Figure S8.

**Fig. 7.**
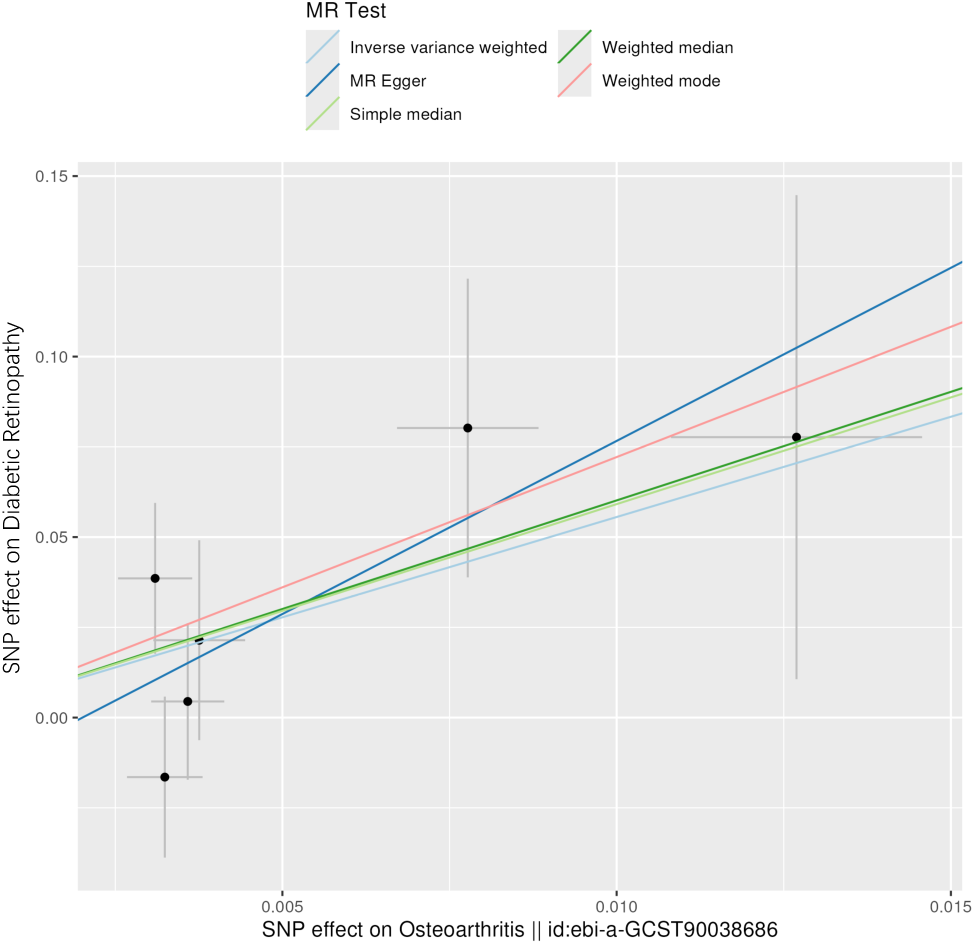
Scatter plot of SNP effects on osteoarthritis (exposure) and diabetic retinopathy (outcome) using multiple MR methods. This scatter plot displays the MR estimates of SNP effects on the relationship between osteoarthritis and diabetic retinopathy using multiple MR methods. The x-axis represents the SNP effects on osteoarthritis, and the y-axis represents the SNP effects on diabetic retinopathy. Different MR methods are indicated by lines of varying colors and styles: inverse variance weighted (green line), weighted median (blue line), MR Egger (red line), simple median (black line), and weighted mode (light blue line). Each point represents a specific SNP, with error bars showing confidence intervals.

## Discussion

In this GWAS study of DR on type 1 patients using the UKB dataset, we identified novel significant genetic variants at a genomic risk locus in the region of gene *CCDC7* and *ITGB1*, highlighting the complex genetic basis associated with DR. The top variants rs184619214 in *CCDC7* and rs79853754 in *ITGB1* have reached genome-wide significance (*p* < 5×10^−8^), and they exhibit high linkage disequilibrium with r^2^ = 0.95. The estimated genomic inflation factor (λ) calculated from GWAS was 0.997 (Fig.4), suggesting a homogeneous population. Notably the heritability of DR was calculated to be 31.09% in the GWAS, emphasizing that DR exhibits a relatively high heritability in the population with T1D.

The rs184619214 variant is located in the *CCDC7* gene (Coiled-Coil Domain Containing 7). Its gene synonyms include *BIOT2*, *C10ORF68*, *FLJ13031*, and *FLJ32762*. Current research on *CCDC7* primarily focuses on its roles in vascular functions and male-specific characteristics. *CCDC7* is involved in VEGF activation to promote sustained angiogenesis, along with many *CCDC* proteins that play a role in vascular stability and inflammation ^31^. According to Priyanka ^32^, *CCDC* proteins are primarily expressed in the human male reproductive system, similar to our gene expression heatmaps showing that *CCDC7* has relatively high expression levels in the testes, suggesting specificity in its function and activity within this system. Research indicates that male sex is a risk factor for DR, with men more prone to advanced retinopathy in T1D and seeming to exhibit more severe conditions at diagnosis in T2D ^33,34^. Conversely, experimental models in mice show no gender differences in the early stages of DR ^35^. In our study according to Table 1 clinical characteristics, there is no significant difference in sex distribution between cases and controls, with a ratio of approximately 57% male to 43% female. This finding indicates that our current study does not provide sufficient evidence to suggest a higher prevalence of DR in males compared to females within T1D. Furthermore, the total sample size of 1,004 may also limit the statistical power of these results.

The rs79853754 variant is located in *ITGB1* (Integrin Subunit Beta 1), which has been found to be associated with DR in previous studies. The *ITGB1* plays a pivotal role in PDR and proliferative vitreo-retinopathy due to its contribution to the terminal event of tractional retinal detachment ^36^. A result suggested a unique mechanism through which *SPARC-ITGB1* interaction and hyperglycemia modulate VEGF signaling to inhibit neovascularization in DR ^37^. Additionally, lysine acetyltransferase 1 (KAT1) induces YTHDF2-mediated instability of *ITGB1* mRNA, which alleviates the progression of DR in a mouse model ^37^. Reactome pathway analysis reveals that *L1CAM* is involved in the assembly of integrin complexes, with *CHL1* mediating interactions between *ITGB1* and *NRP1*. Elevated levels of *L1CAM* have been associated with retinopathy ^38^. Additionally, there is a partial overlap between the *ITGB1* and *NRP1* genes, with *NRP1* identified as being linked to DR ^39,40^. Inhibition of *NRP1* has been shown to reduce the development of early DR ^41^. Furthermore, under hypoxic conditions, *CHL1* upregulation disrupts the homeostatic balance between BMP-4 and VEGF, enhancing VEGF’s role in promoting retinal angiogenesis ^42^. Other comprehensive analyses using tissue expression analysis, TWAS, and eQTL studies across multiple tissue types from the GTEx database did not yield significant findings for our significant SNPs. This suggests that while *ITGB1* is known to play a crucial role at the cellular level, there is no evidence from our data to indicate that these SNPs have tissue-specific expression patterns.

Gene expression heatmaps indicate higher expression in both the artery and nerve tibial regions (Fig.5). Research links tibial nerve characteristics to DR. patients with mild NPDR show reduced tibial motor conduction velocity compared to those without DR ^43^. Moreover, diabetic peripheral neuropathy (DPN) patients exhibit increased cross-sectional areas of the tibial nerve, while DPN and DR have a positive correlation ^44^, with similar abnormalities noted in type 1 diabetes (T1D) patients with neuropathy ^45^, suggesting the tibial nerve’s involvement in both T1D and DR. It is noteworthy that *CCDC7* has been linked to tibial involvement, as it was identified in patients with symptoms similar to centronuclear myopathy, featuring lower limb pain and reduced muscle strength in the tibia ^46^. Chromatin interaction analysis revealed significant interactions in *CCDC7* and *ITGB1*, where common cell and tissue are Mesenchymal Stem Cell and Left Ventricle. Mesenchymal stem cells have been discovered as potential therapy in diabetic retinopathy ^47,48^, and left ventricle structure and dysfunction were associated with DR^49^.

The PheWAS results have provided us with many important insights. Eotaxin (CCL11) is a chemokine closely associated with inflammatory processes and has been recognized as a representative phenotype indicating the presence of inflammation in the body. Furthermore, Eotaxin (CCL11) is proposed as a potential candidate for developing targeted therapeutic strategies, which could be utilized either alone or in conjunction with anti-VEGF drugs to treat PDR ^50^. Radiation retinopathy is more likely to develop in eyes with preexisting diabetic retinopathy and, due to their clinical and histopathological similarities, is treated using established diabetic retinopathy therapies like laser photocoagulation ^51,52^. M23 Internal derangement of the knee and osteoarthritis of the hip from the skeletal domain showed significant associations with most of the SNPs. Research indicates that PDR is significantly associated with the absence of clinical OA in long-term diabetes mellitus patients ^53^. Other studies found that retinal arteriolar narrowing, associated with an increased risk of knee replacement for OA ^54^, also correlated with diabetic retinopathy in T1D patients ^55^. Our MR analysis demonstrates that OA acts as a significant exposure factor associated with the development of DR. This suggests that OA may play a mechanistic role in the pathogenesis of DR, potentially mediated by systemic inflammatory processes or microvascular dysfunction that are common to both conditions. Identifying OA as a contributory factor to DR provides a valuable early warning indicator, which could be critical for preemptive medical intervention to mitigate the progression of DR.

Our study’s limitation stems from the UKB’s unclear definition of T1D. We established our T1D definition based on clinical insights, considering both the sample size (a minimum of 1000 participants) and the precision (age at diabetes diagnosis) to finalize our definition of T1D (Fig.1). Compared to another study’s definition of T1D—diagnosed under 30 years and requiring insulin within a year of diagnosis ^19^—our clinical characteristics showed an average age at diabetes diagnosis of approximately 22 years. Furthermore, unlike Forrest ^56^, which included diverse ethnic backgrounds from the UKB cohort, our study targeted White British individuals with T1D-related DR. We also incorporated critical covariates such as DoD and HbA1c levels in our GWAS, enhancing both the accuracy and relevance of our findings. Our significant SNPs have proven challenging to replicate strongly. This may be attributed to the limited number of studies focusing on this specific association, as detailed in Supplementary Table S7, resulting in insufficient robust data to support our findings. Although the meta-analysis indicates significant results, the use of high linkage disequilibrium proxies for these SNPs introduces a potential bias and limits the statistical power of our analyses.

## Conclusion

Overall, our GWAS study identified a new locus at the genetic position of the *CCDC7* and *ITGB1* genes, which will help to understand the mechanisms of DR.

## Statements and Declarations

### Data availability

This research complies with all ethical standards and data privacy measures the UKB sets. Detailed summary statistics data about DR within the UKB are retrievable from https://figshare.com/s/68c2e17003c694c6f453. Should any additional data pertinent to this study need to be presented within this paper or its other files, the authors can provide such data upon reasonable request.

#### Author information

### Authors and Affiliations

**Nottingham Ningbo China Beacons of Excellence Research and Innovation Institute, University of Nottingham Ningbo China, Ningbo, China, 315100**

Tengda Cai, Qi Pan, Yiwen Tao, Luning Yang, Weihua Meng

**Division of Population Health and Genomics, School of Medicine, University of Dundee, Dundee, UK, DD2 4BF**

Charvi Nangia, Colin NA Palmer

**Biodemography of Aging Research Unit, Duke University, Durham, NC, USA, 27708-0408**

Aravind L Rajendrakumar

**NIHR Global Health Research Unit on Global Diabetes Outcomes Research, Division of Population Health and Genomics, University of Dundee, Ninewells Hospital and Medical School, Dundee, UK, DD1 9SY**

Yu Huang

**Department of Ophthalmology, The Affiliated Ningbo Eye Hospital of Wenzhou Medical University, Ningbo, China. 315040**

Yongqing Shao

**Department of Ophthalmology, Lihuili Hospital affiliated with Ningbo University, Yinzhou District, Ningbo, China**

Yunyan Ye

**School of Veterinary Medicine and Science, University of Nottingham, Nottingham, UK, LE12 5RD**

Tania Dottorini

**School of Mathematical Sciences, University of Nottingham Ningbo China, Ningbo, China, 315100**

Mainul Haque

## Authors’ Contributions

TC drafted the paper and performed GWAS analysis. QP, YT, LY contributed to data formatting and correction. CN, AR, YH, YS, YY, TD, MH, CP provided comments on the paper. WM organized the project and provided comments.

## Acknowledgment

This study adheres to all ethical guidelines and data protection protocols of the UK Biobank. The current study was conducted under approved UK Biobank data application number 50604. This study was mainly funded by the Pioneer and Leading Goose R&D Program of Zhejiang Province 2023 with reference number 2023C04049 and Ningbo International Collaboration Program 2023 with reference number 2023H025. The authors gratefully thank all the participants and professionals contributing to the UKB and FinnGen studies.

## Corresponding authors

Correspondence to Weihua Meng

## Consent to Publish

All authors have consent for publication.

## Ethics approval

This study was approved by the Ethics Committee of the University of Nottingham, Ningbo, China.

## Figure legends

**Figure S1.** (A) The forest plots of the meta-analyses results from GWAS conducted by UKB study with European, FinnGen, GoSHARE, GoDARTS and Caucasian Australian study. Lines with green colors represent the effect sizes of SNPs from different cohorts. Each green line corresponds to a specific cohort, indicating the estimated effect size along with its 95% confidence interval. The overall effect beta (with CIs) is summarized at the bottom of the plot, shown as a red diamond. (B) Funnel plot of meta-analysis results. The horizontal axis represents the effect estimates beta, and the vertical axis denotes the standard error of each study included in the meta-analysis. The funnel plot is primarily symmetric around the vertical dashed line, which represents the overall effect estimate. This symmetry suggests a minimal risk of publication bias within the included studies. (C) Radial plot of meta-analysis results. Displaying the influence of individual studies on the overall effect estimate. The horizontal axis represents the square root of the inverse of the variance, which corresponds to the precision of each study. The vertical axis shows the standardized effect sizes. The grey shaded area delineates the 95% confidence region around the combined effect estimate, illustrating the consistency of individual study results with the overall meta-analytic conclusion.

**Figure S2.** The Manhattan plot of gene analysis in the primary GWAS. Genome-wide significance was defined and established after Bonferroni correction (p = 0.05/19295 = 2.59 x10-6), as illustrated by the red dashed line in the plot.

**Figure S3.** Tissue expression results on 30 general tissue types and 53 specific tissue types by GTEx in the FUMA. The dashed line shows the cut-off *p* value for significance with Bonferroni adjustment for multiple hypothesis testing. No tissues achieved statistical significance

**Figure S4.** Tissue Specificity of Differentially Expressed Genes in GTEx v8 Data. The figure presents bar plots illustrating the results of an enrichment test for differentially expressed genes (DEGs) across two datasets: GTEx v8 with 54 specific tissue types (left panel) and GTEx v8 with 30 general tissue types (right panel). Each bar represents the –log10 transformed *p* values of DEGs tested for enrichment in positional mapping genes within each tissue type. Significant enrichment is highlighted in the brain cortex in the 54 tissue type dataset, shown by the red bar, indicating a notable down-regulation of DEGs. This analysis focuses on the enrichment of positional mapping genes and avoids the broader SNP p-value distribution typically used in MAGMA tissue expression analyses.

**Figure S5.** Enrichment Analysis of Genetic Associations. This figure presents the results of enrichment analyses for positional gene sets (MsigDB c1), Reactome pathways (MsigDB c2), and GWAS catalog reported genes related to specific traits. The analysis reveals varying degrees of gene overlap and enrichment significance across different datasets. The proportion of overlapping genes and their enrichment p-values are displayed for each category, highlighting notable interactions in biological pathways.

**Figure S6.** Gene Association Patterns Across Human Tissues from MSigDB Chr10p11. This heatmap depicts the expression and association patterns of multiple genes across a variety of human tissues, as sourced from the Novartis Human Tissue Compendium. Notably, the genes *ITGB1* and *NRP1* are located under the same detailed node in the tree diagram.

**Figure S7.** Pathway Diagram of *ITGB1* from Reactome. (A) showed interaction pathways involving *L1CAM*, *ITGB1*, and *NRP1*. *L1CAM* is shown to participate in the assembly of integrin complexes. (B) depicts *CHL1*’s role in facilitating interactions between *ITGB1* and *NRP1*.

**Figure S8.** Mendelian Randomization Analysis between Osteoarthritis and Diabetic Retinopathy. Utilizing genetic variants as instrumental variables to estimate the causal effects between OA and DR. The first plot (top panel) shows the forest plot, funnel plot, and leave-one-out plot of MR results when osteoarthritis is considered the exposure and diabetic retinopathy the outcome. The second plot (bottom panel) reverses the roles, treating DR as the exposure and OA as the outcome. The plot of SNP effects using multiple MR methods is shown, as well as the forest plot, funnel plot, and leave-one-out plot.

