## Supplementary figures and images for "Genome-wide association studies found *CCDC7* and *ITGB1* associated with diabetic retinopathy"

### Figure S1

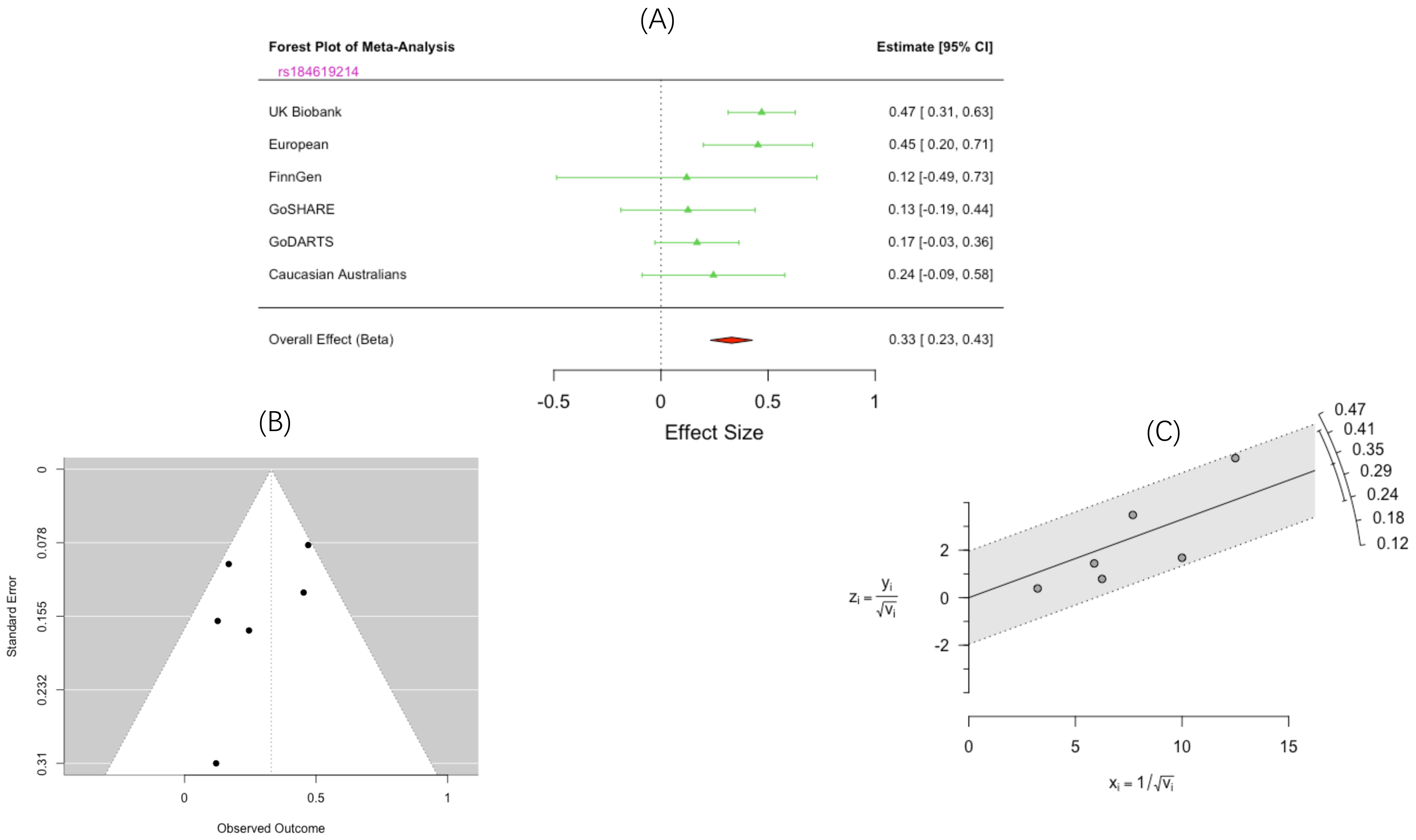

### Figure S2

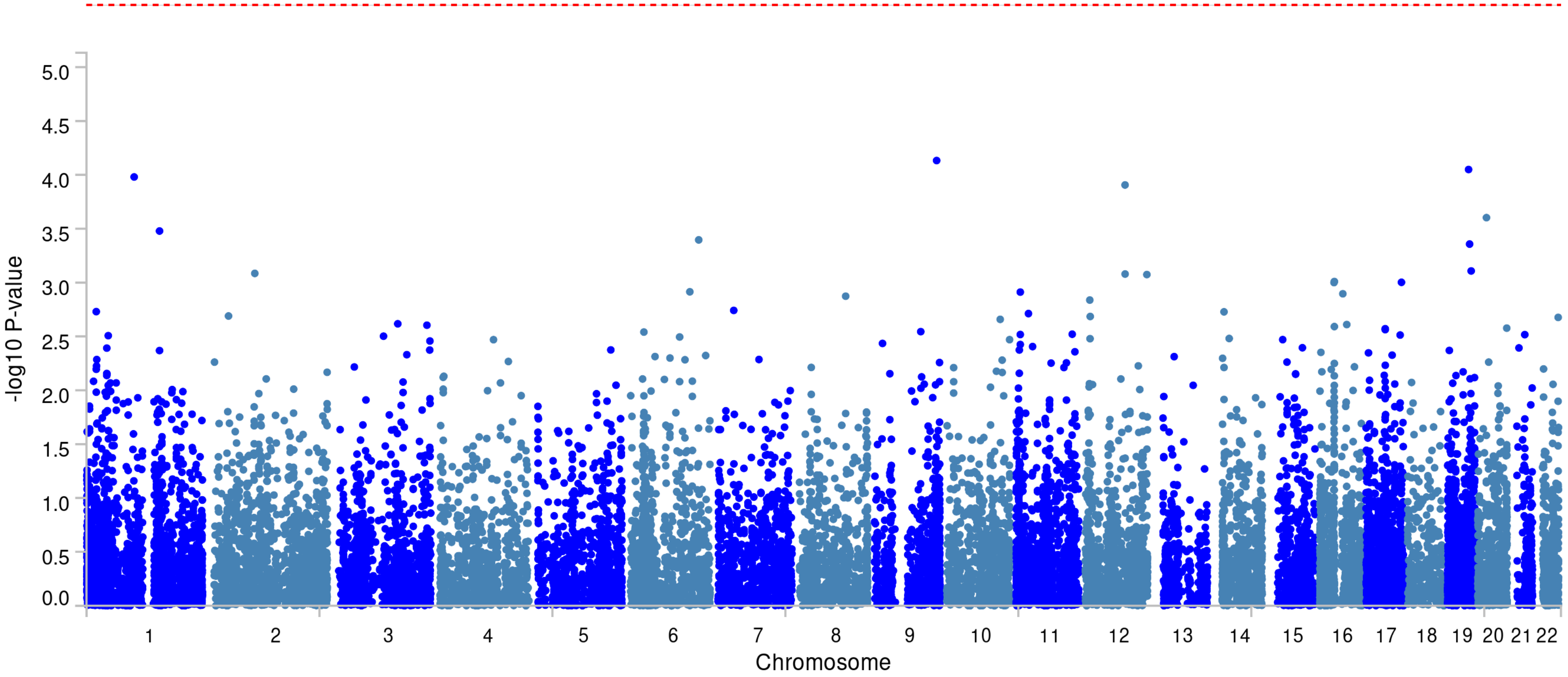

### Figure S3

#### MAGMA tissue expression analysis

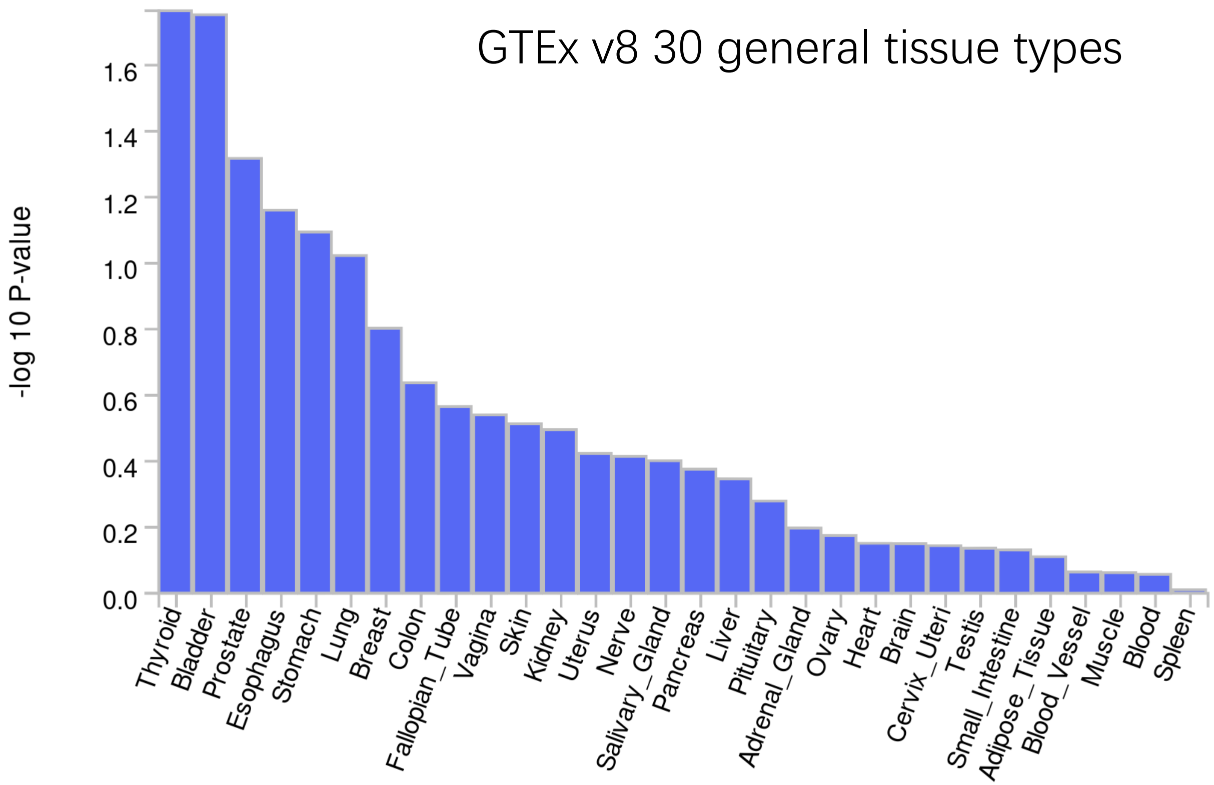


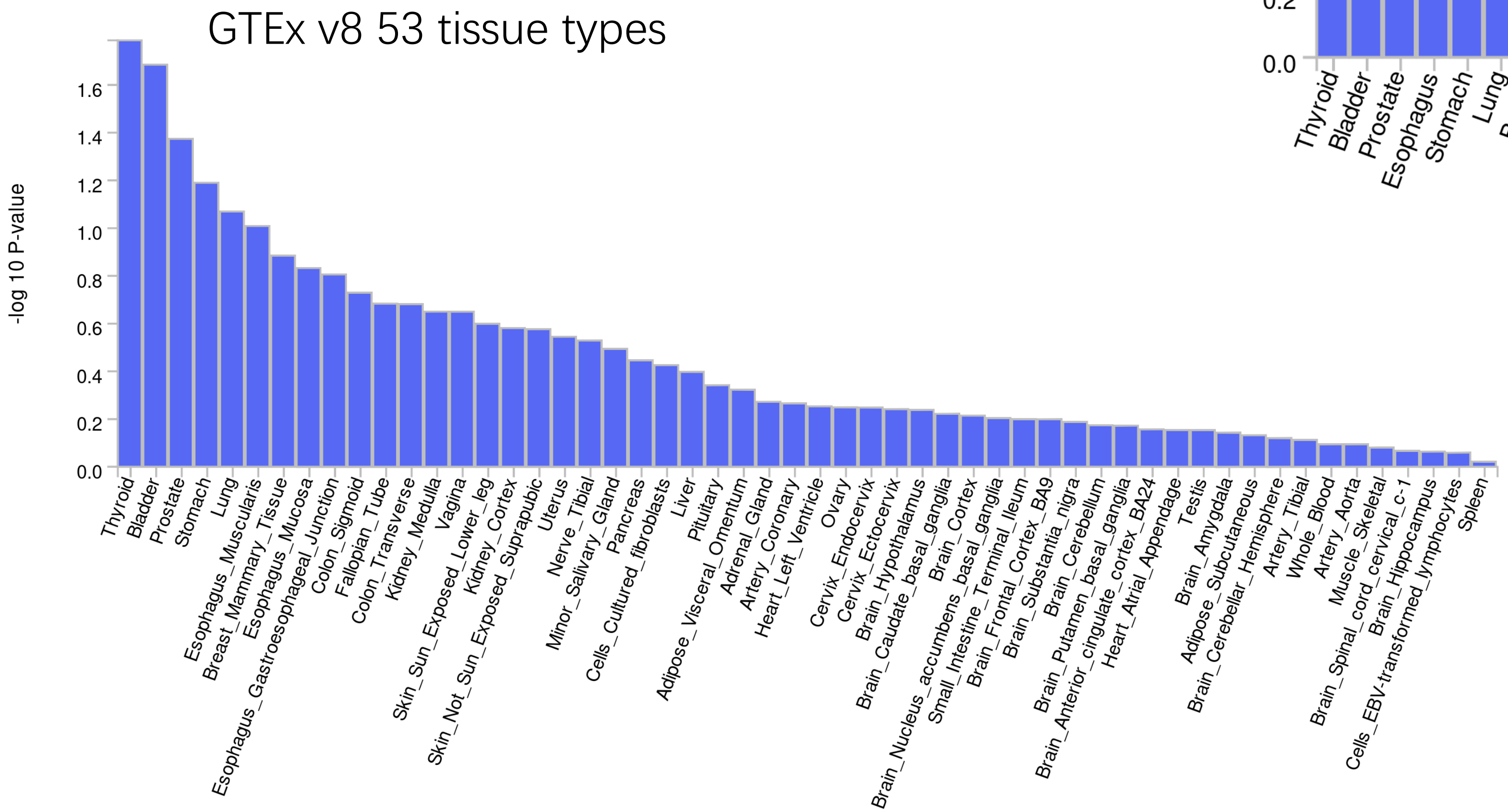

### Figure S4

##### GTEx v8 54 general tissue types

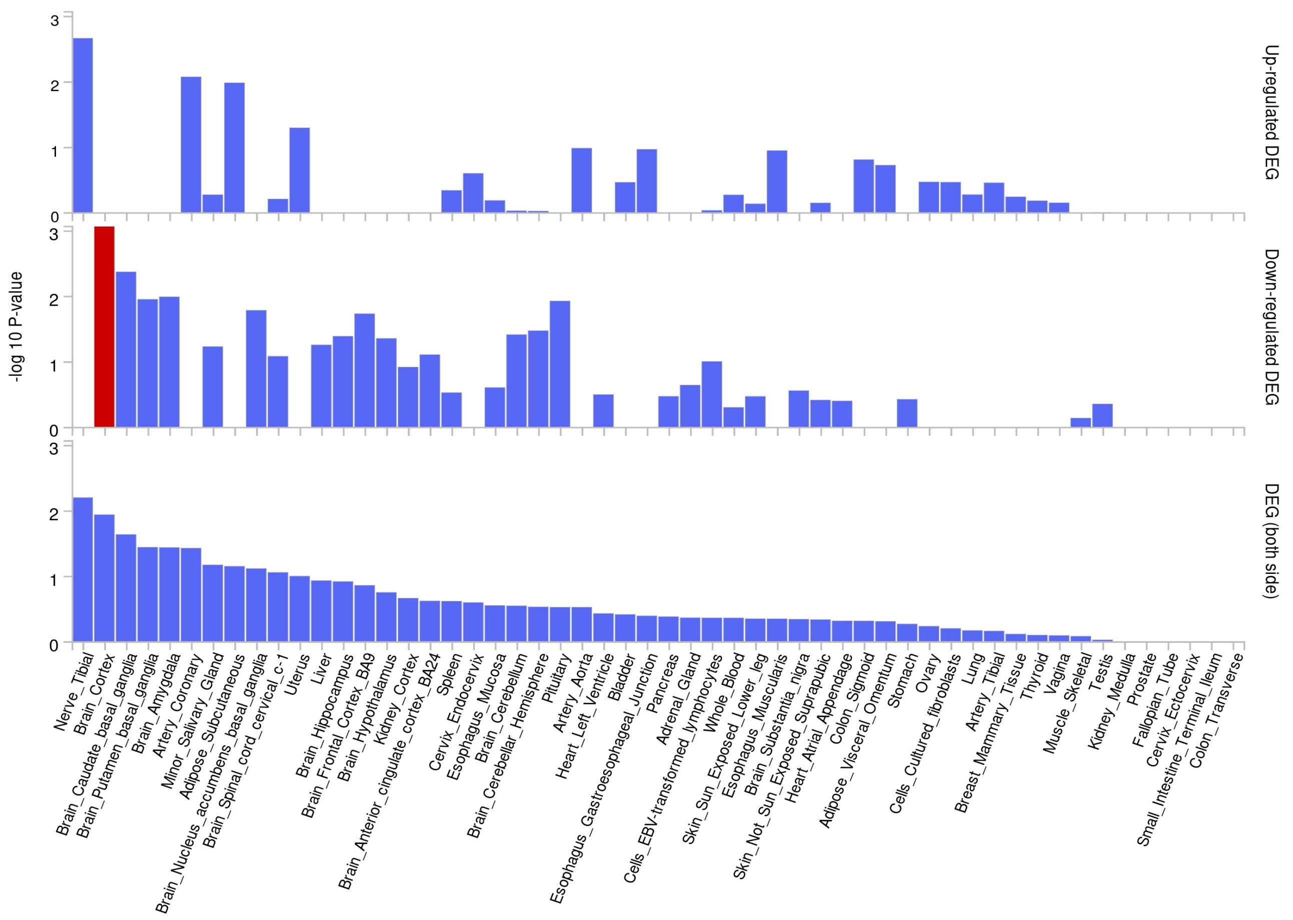


##### GTEx v8 30 general tissue types

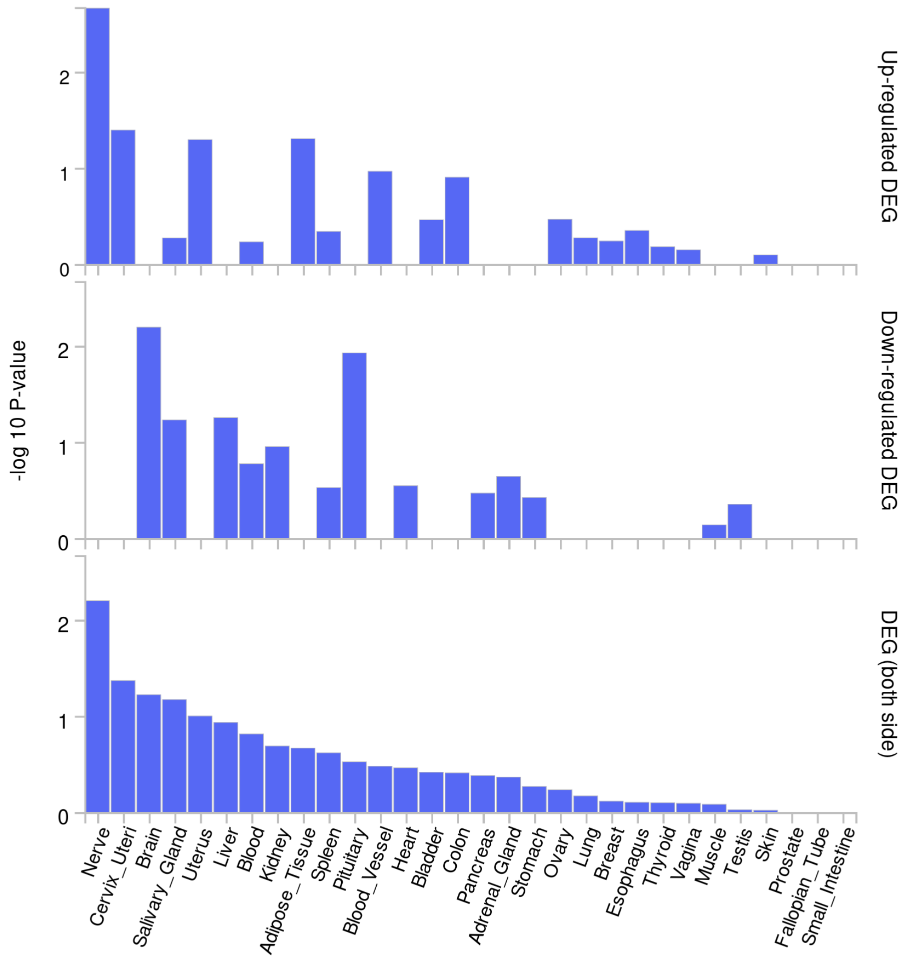

### Figure S5

Positional gene sets (MsigDB c1)

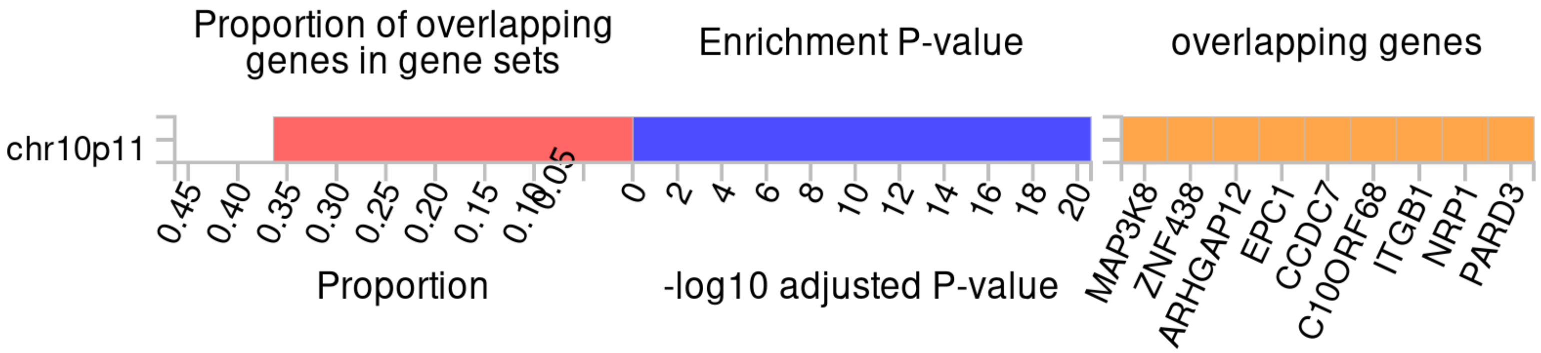


Reactome (MsigDB c2)

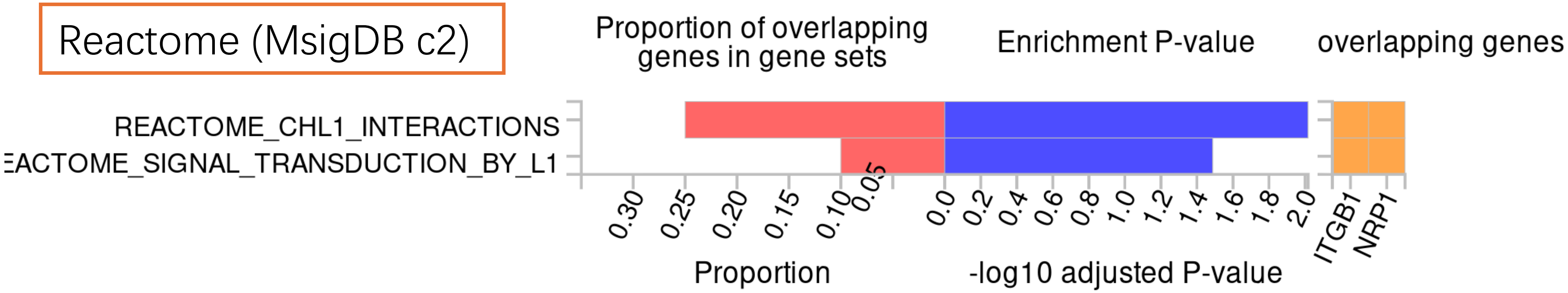


GWAS catalog reported genes

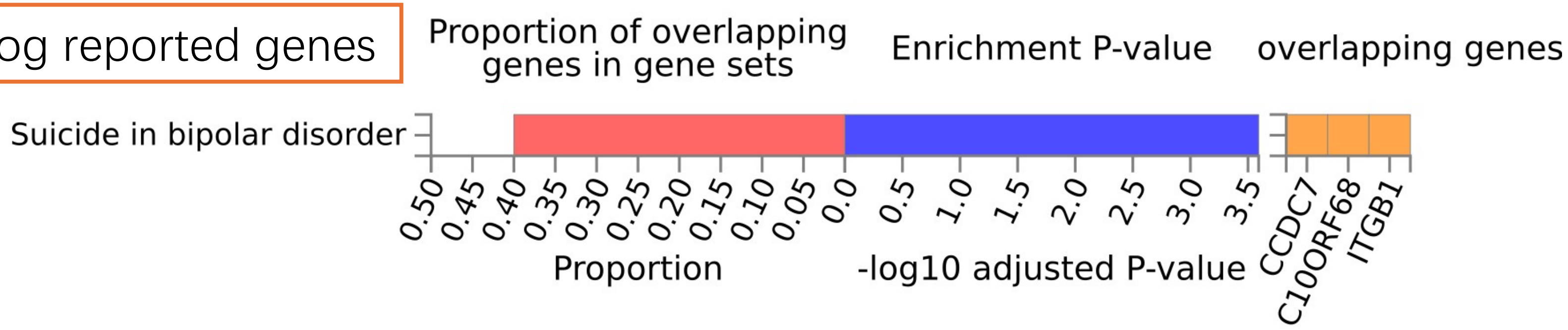

### Figure S6

#### Human tissue compendium (Novartis)

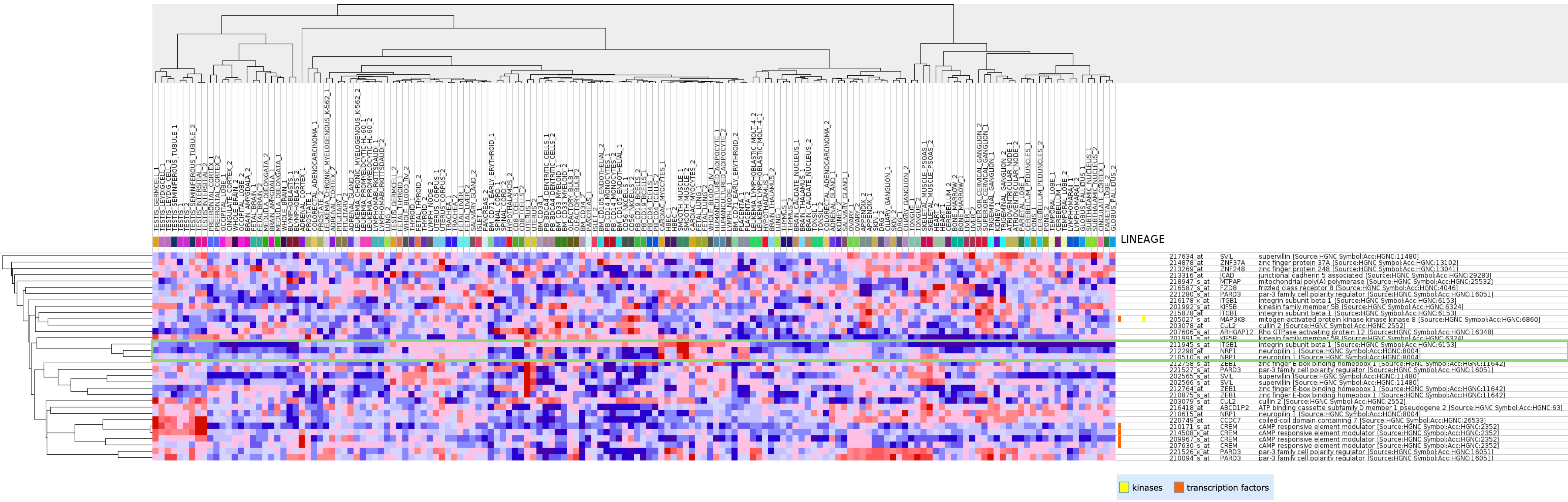

### Figure S7

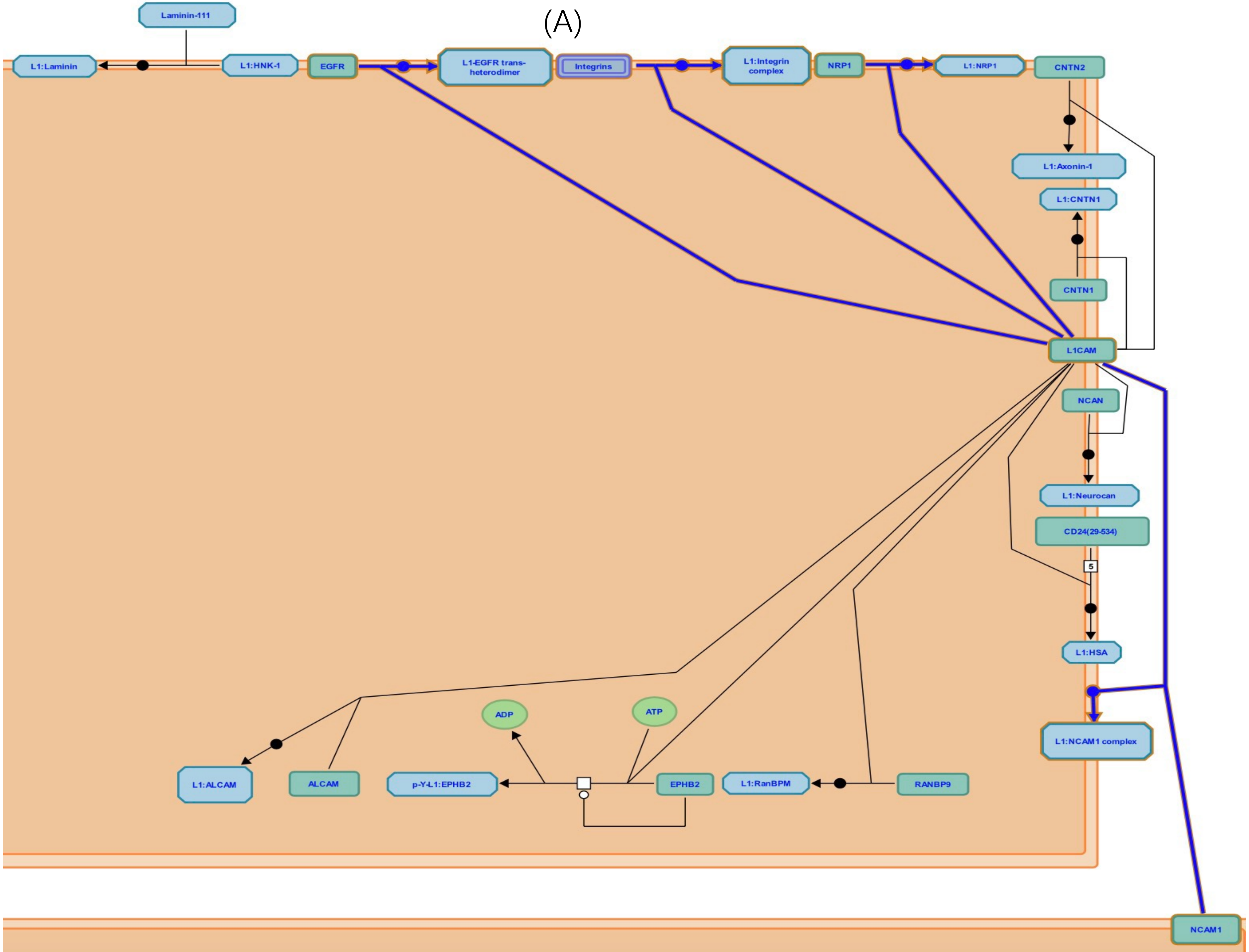

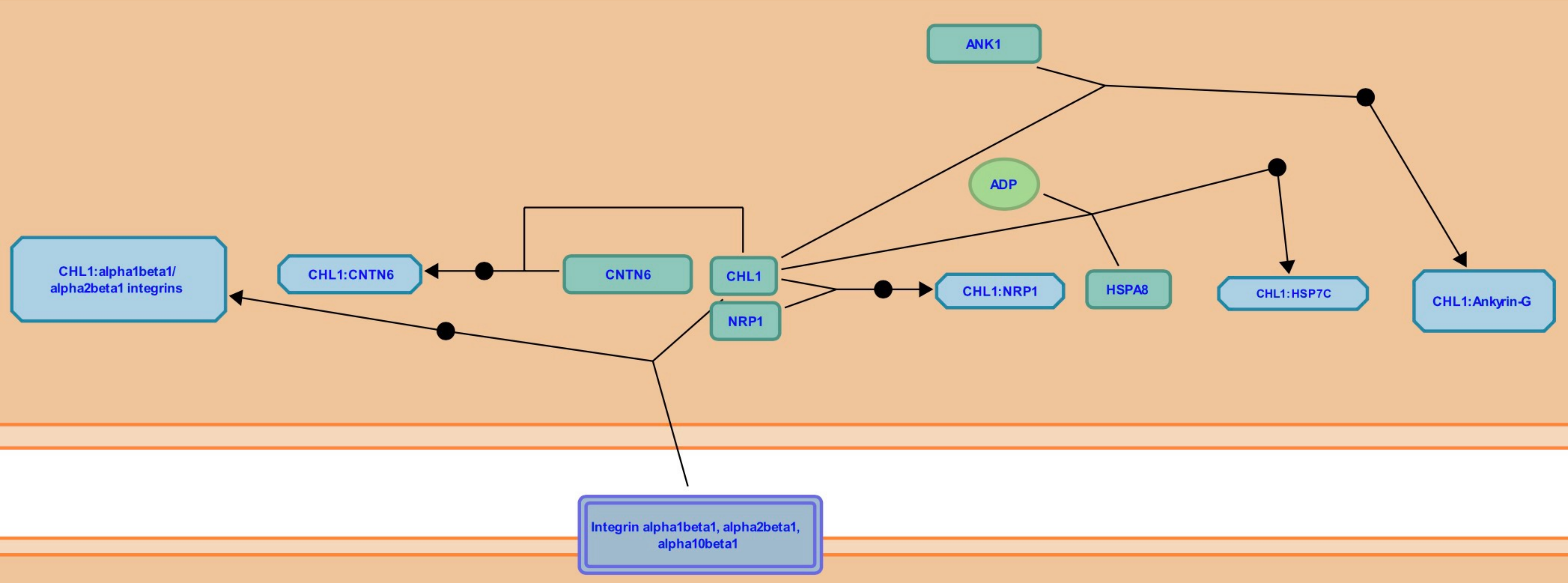
