## Supplementary material for "Genome-wide association studies found *CCDC7* and *ITGB1* associated with diabetic retinopathy": Figure S8

Mendelian Randomization

### Osteoarthritis (exposure), Diabetic retinopathy (outcome)

#### Forest plot

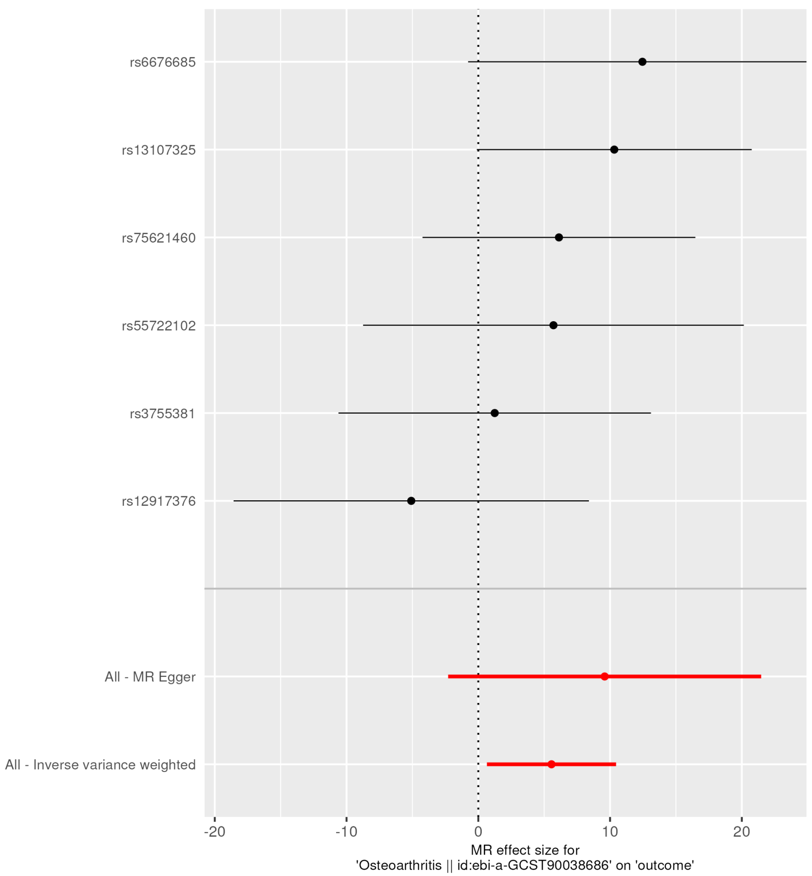


#### Funnel plot

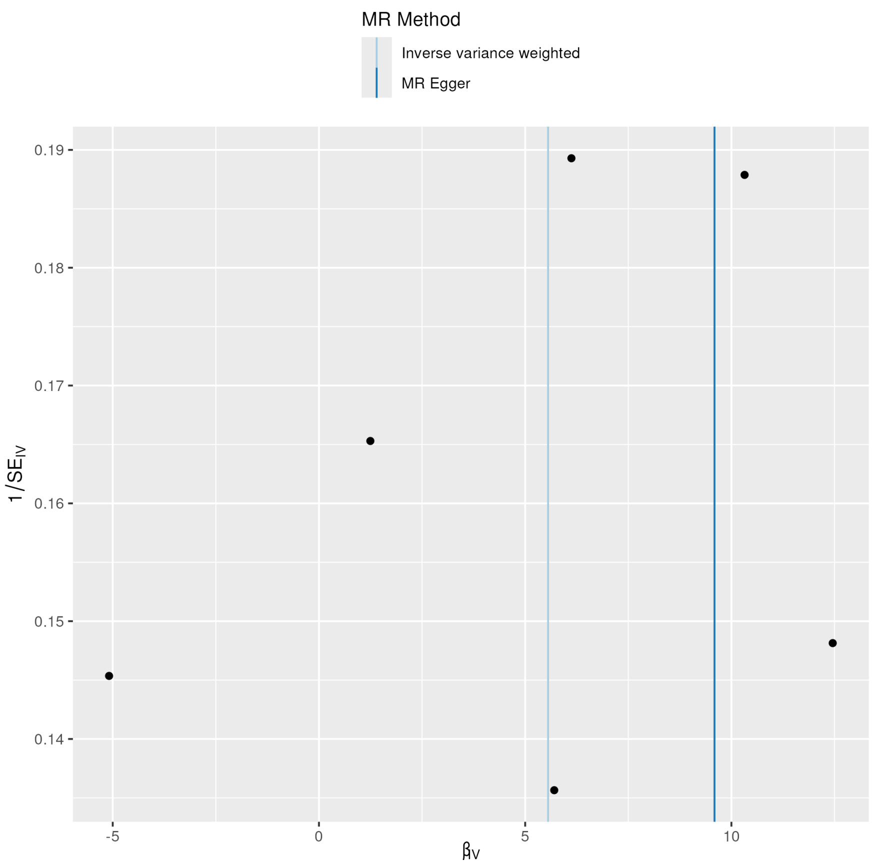


#### Leave-one-out plot

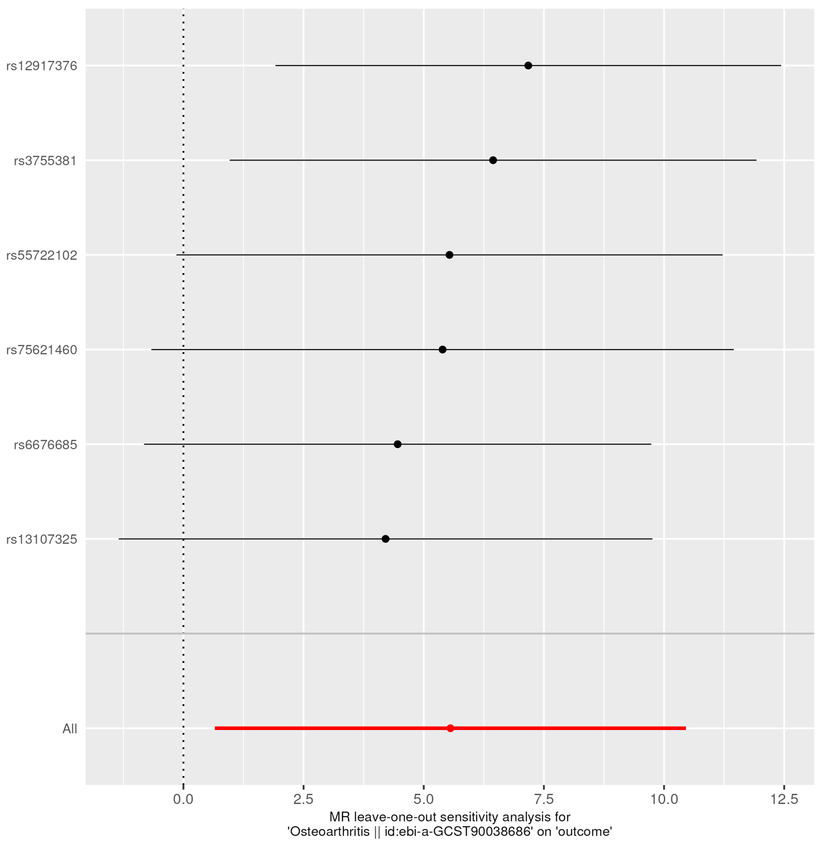

### Mendelian Randomization

Diabetic retinopathy (exposure), Osteoarthritis (outcome)

MR_results plot

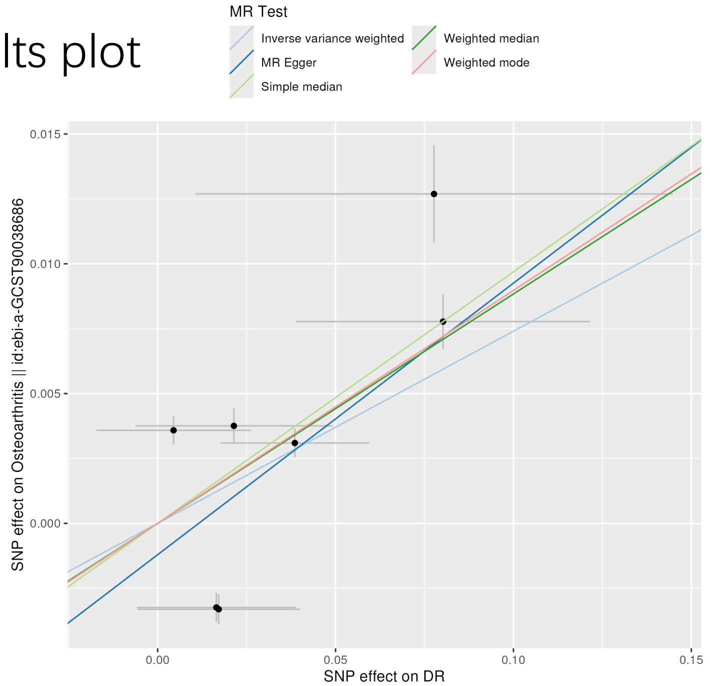


Forest plot

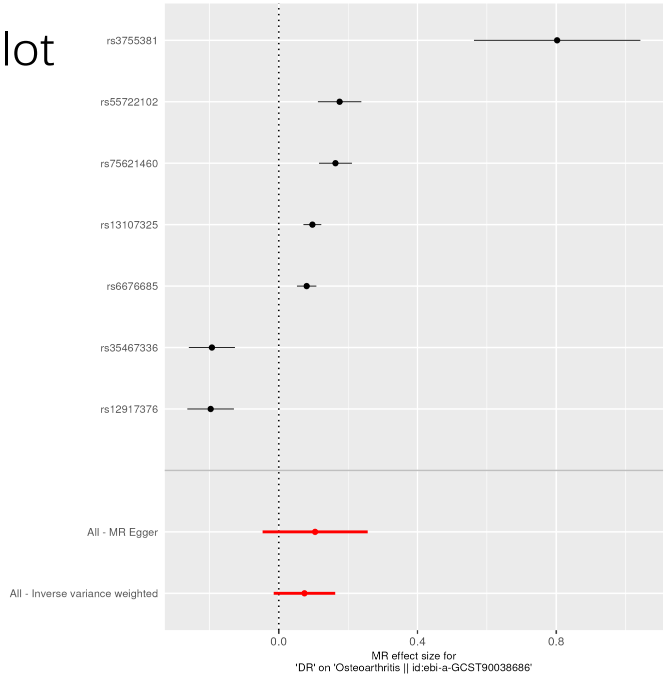


Funnel plot

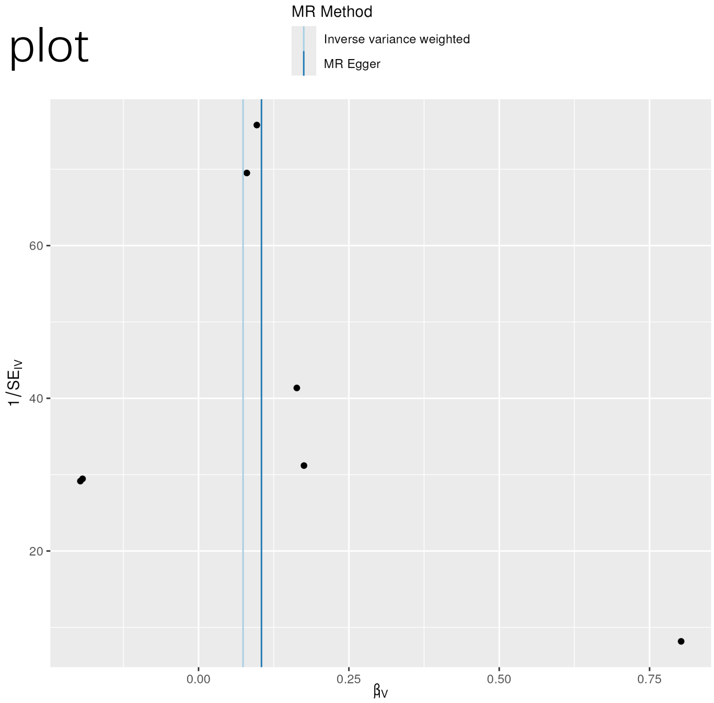


Leave_one_out plot

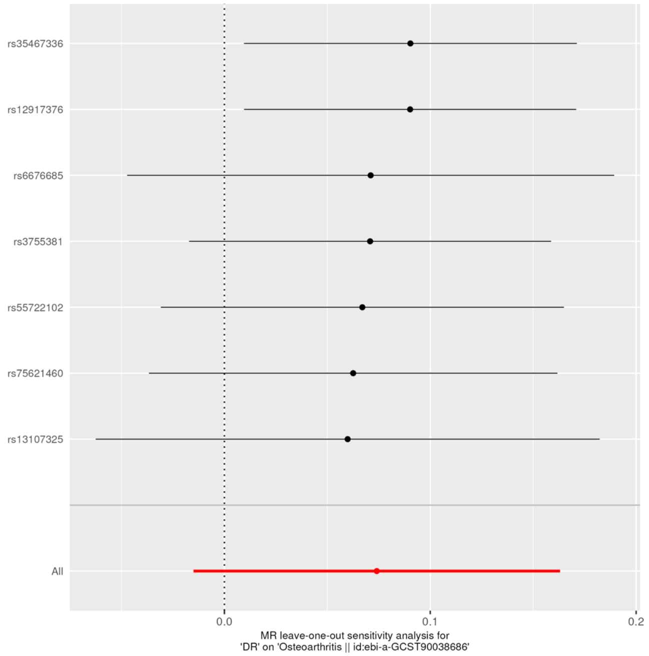
